# Digital access, media exposure, and maternal health service utilization among women in Ghana

**DOI:** 10.64898/2026.09.15.26363143

**Authors:** Sandy Baffour Adu, Nana Akwasi Owusu Ahenkan, Jochebed Afua Basil, Kelvin Akoto Gyamera, Sophia Ofosua Ansong, Yaw Ofosu Ansong Snr

**Affiliations:** Maternal Health Research Division, Summer Health Limited, Accra, Greater Accra, Ghana; Department of Obstetrics and Gynaecology, University of Cape Coast School of Medical Sciences, Cape Coast, Ghana; Department of Obstetrics and Gynaecology, Lincoln County Hospital, Greetwell Road, United Kingdom; Department for Continuing Education, 1 Wellington Square, University of Oxford, Oxford, United Kingdom

## Abstract

Maternal health service utilization remains a critical strategy for reducing maternal and neonatal mortality, yet gaps persist across antenatal care (ANC), skilled birth attendance (SBA), and postnatal care (PNC) in Ghana, particularly among rural women. While mass media and digital access have been linked to improved utilization of maternal health services in Sub-Saharan Africa, evidence specific to Ghana remains limited. This study assessed the association between media exposure (radio, television, newspaper) and digital access (phone ownership, internet use) with four maternal health service utilization outcomes: ANC use (4+ visits), adequate ANC (8+ visits), SBA, and PNC among women in Ghana, stratified by place of residence. We analyzed data from 4,982 women in the 2022 Ghana Demographic and Health Survey using multilevel logistic regression, adjusting for sociodemographic and healthcare-access covariates. Utilization was high for ANC4+ (88.5%), SBA (86.7%), and PNC (86.3%), but substantially lower for ANC8+ (38.7%), with rural women underperforming urban women across all outcomes. After adjustment, mobile phone ownership emerged as the most consistent correlate of service utilization. Basic phone ownership was associated with higher odds of ANC use, SBA, and PNC, while smartphone ownership was associated with higher odds of ANC use and SBA. Weekly radio listening was associated with higher odds of adequate ANC in the total sample, but no other media exposure variable or regular internet use was independently associated with any outcome. These findings suggest that mobile phone ownership is more consistently associated with maternal healthcare use in Ghana than traditional mass media exposure or internet use. Health policy and implementation research is therefore needed to examine how mobile phone-based approaches, differentiated by phone type and tailored to urban and rural contexts, may support efforts to reduce persistent disparities in maternal health service utilization in Ghana.

## Introduction

Maternal mortality in Ghana remains a major public health concern, with rural and low-income women disproportionately affected despite Ghana’s gradual progress toward global health targets [1–3]. Existing estimates from 2023 indicate that Ghana’s maternal mortality rate was 234 per 100,000 live births, which is considerably lower than the Sub-Saharan African (SSA) average (448 per 100,000 live births) but above the average for Lower Middle-Income countries (178 per 100,000 live births) and the global average of 197 [4]. Addressing this challenge requires targeted national interventions aimed at improving access to and utilization of maternal health care in Ghana, as substantial evidence has shown that the use of maternal health services is a key strategy for preventing pregnancy-related complications and reducing maternal and neonatal mortality [5–9].

Maternal health services comprise antenatal care (ANC), intrapartum care provided by skilled birth attendants (SBA), and postnatal care (PNC), which are recognized as key determinants of reduced maternal and neonatal mortality [10]. However, major gaps in antenatal care, skilled birth attendance, and postnatal care continue to undermine maternal and neonatal health outcomes across Ghana and Sub-Saharan Africa [11]. For instance, an analysis of Demographic and Health Survey (DHS) data from 26 SSA countries found that women with at least one ANC visit were 48% less likely to experience neonatal death, mortality was 2.2-fold higher among those who received no ANC. Yet only 58.4% completed four or more ANC visits, highlighting persistent barriers to continuity of maternal care and progress toward the maternal and neonatal targets of the Sustainable Development Goals (SDGs) [12].

Traditional mass media, including radio, television, and newspapers, alongside emerging digital technologies such as mobile phones and the internet, provide important platforms for disseminating maternal health information, promoting healthy behaviours, facilitating communication with healthcare providers, and encouraging timely utilization of maternal healthcare services [13–15]. Evidence from low- and middle-income countries (LMICs) indicates that mass media exposure and digital access are associated with greater maternal health service utilization [13,14,16,17]. A study conducted across 28 SSA countries, including Ghana, reports that women who listen to radio and watch television at least once a week are more likely to utilize ANC and SBA [18]. Another study, which investigated women in Nepal, Liberia, Senegal, Nigeria, and Rwanda, provided strong support for the argument that access to and use of digital resources are strongly associated with health, though this varies by context and specific health outcome [19]. Together, these findings suggest that media and digital platforms may support utilization of maternal health services by increasing exposure to maternal health information and facilitating engagement with healthcare services.

Although considerable research has examined associations between mass media exposure, digital access, and maternal health service utilization, these relationships remain inadequately characterized in Ghana. To the best of our knowledge, Ghanaian-specific studies examining associations between media exposure, digital access, and maternal health service utilization have been limited to either skilled birth attendance or health facility delivery [13,14,17], rather than the broader perinatal care continuum.

Therefore, this study aims to assess the association between different forms of mass media exposure, digital access, and utilization of maternal health services among women in Ghana, stratified by place of residence. The findings could help fill an important evidence gap in maternal health service utilization in Ghana, particularly among rural women. They may also identify the forms of mass media and digital access associated with improved utilization of maternal health services in Ghana, thereby informing interventions to reduce maternal and neonatal mortality.

## Methods

### Study design and data source

This study utilized data from the 2022 Ghana Demographic and Health Survey (GDHS), a nationally representative cross-sectional survey conducted by the Ghana Statistical Service in collaboration with the Ghana Health Service between November 2022 and March 2023. The survey used a cross-sectional design with a two-stage stratified cluster sampling technique. Enumeration areas were selected in the first stage using probability proportional to size sampling, and households were systematically selected in the second stage. The data were extracted from the women’s Individual Recode file of the 2022 GDHS, which contains one record per woman aged 15-49 years. The analytical sample included 4,982 women at the individual level and 617 clusters at the cluster level. The dataset used in this study is publicly available at https://dhsprogram.com and was accessed with permission from the DHS Program following registration “S1 File”. As a secondary analysis of anonymized, publicly available data, this study did not require separate ethical review.

### Variables

#### Outcome variables

Maternal health service utilization was assessed using four outcomes: ANC4+, ANC8+, skilled birth attendance (SBA), and postnatal care (PNC). For adequate ANC (ANC8+), women were asked how many antenatal visits they had during their most recent pregnancy. Responses were recorded as a count variable and recoded as no adequate ANC (0 to 7 visits = 0) or adequate ANC (8 or more visits = 1), reflecting the WHO 2016 recommendation of a minimum of eight antenatal contacts during pregnancy [20]. For ANC use, the same variable was recoded as no ANC4+ use (0 to 3 visits = 0) or ANC4+ use (4 or more visits = 1). ANC4+ remains a widely used indicator in DHS-based maternal health studies and allows comparison with prior literature [13,21]. For skilled birth attendance (SBA), women who received delivery assistance from a doctor or nurse/midwife were coded as yes (= 1); women not meeting this criterion were coded as no (= 0) [22]. For postnatal care within six weeks (PNC), women who received a postnatal check from a doctor or nurse/midwife within six weeks of delivery were coded as yes (= 1); women not meeting this definition were coded as no (= 0) [23].

#### Exposure variables

The main exposures were media exposure (radio, television, and newspaper) and digital access (mobile phone ownership and internet use). Media exposure was assessed using frequency of radio listening, television viewing, and newspaper reading; each variable was categorized as not at all, less than once a week, at least once a week, or almost every day. The almost every day category had zero observations for all three media variables in the Ghana 2022 dataset; the final analytic categories were therefore coded as 0 = not at all, 1 = less than once a week, and 2 = at least once a week [18]. For digital access, phone type was constructed from ownership of any mobile phone (V169A) and smartphone ownership (V169C), and coded as 0 = no phone, 1 = basic phone only, and 2 = smartphone. Internet use was derived from frequency of internet use in the last month (V171B) and dichotomized as 0 = not a regular user, defined as not at all or less than once a week, and 1 = regular user, defined as at least once a week or almost every day [24].

#### Covariates

The covariates were selected based on their established association with the outcome variables in the literature [24–26] and their theoretical relevance as guided by the Andersen Behavioural Model of Health Services Use [27]. Age group was retained in its original seven-category form (15 to 19 through 45 to 49 years). Educational level was retained as four categories: no education, primary, secondary, and higher. Religion was recoded into four categories: Christian, Muslim, traditional, and no religion/other. Marital status was recoded as 0 = never married, 1 = married, 2 = cohabiting, and 3 = formerly married; the formerly married category combined widowed, divorced, and separated women because of small cell sizes. Current working status was retained as a binary variable (not working or working). Parity was recoded as none, one, two, three, and four or more. Health insurance status was retained as a binary variable (not insured or insured). Wealth index was retained as the DHS preconstructed five-category composite. Place of residence was coded as a binary variable (rural or urban) and included as a covariate only in total-sample models. Electricity access was coded as a binary variable, with non-de jure residents coded as missing. Region (V024) was entered into all multilevel models as a 16-category factor variable, with Western Region as the reference category because it was the first coded category; the choice of reference region was otherwise arbitrary. A composite healthcare access barrier to variable was constructed from four items: permission to seek care, money for treatment, distance to facility, and not wanting to go alone, and was coded as 1 if any barrier was reported as a big problem and 0 otherwise.

### Statistical analyses

Weighted prevalence estimates with 95% confidence intervals were computed for each outcome across all explanatory variables and covariates using the DHS sampling weight (V005/1,000,000), while accounting for the clustered and stratified survey design. The Rao-Scott adjusted F test was used to assess the statistical significance of bivariate associations. Before regression modelling, multicollinearity among all predictors, including digital access, media exposure, and socioeconomic covariates that might plausibly overlap (wealth index, electricity access, phone ownership, internet use, and educational level), was assessed using variance inflation factors (VIFs), which confirmed no evidence of problematic multicollinearity “S1 Table”.

Multilevel logistic regression was used to examine associations between each exposure domain and each outcome, with women nested within clusters [28]. Three models were fitted sequentially: a random-intercept-only null model (Model 0), an unadjusted model including exposure variables only (Model I), and a fully adjusted model including all covariates (Model II). Media exposure and digital access variables were modeled separately. Adjusted odds ratios (aORs) with 95% confidence intervals are reported alongside the intraclass correlation coefficient (ICC) and median odds ratio (MOR). All models were estimated for the total sample and separately for urban and rural strata.

Missing data on the analytic variables affected 1,983 of 6,965 eligible women (28.5%), predominantly because of missing responses for maternal healthcare utilization questions, particularly ANC, SBA, and PNC variables (26.0% to 27.2% each) “S2 Table”. To assess the robustness of complete case estimates to this missingness, multiple imputation by chained equations (MICE) was conducted using 30 imputed datasets. The findings from the imputed analyses were largely consistent with those from the complete-case analyses, with no substantive changes in the direction or interpretation of the associations, suggesting that the study conclusions were robust to missing data “S3 Table”. All analyses were conducted in Python 3.12 using Google Colaboratory, with multilevel models fitted in R version 4.x via the lme4 package.

## Results

### Sociodemographic and reproductive characteristics of women

“Table 1” presents the background characteristics of the analytic sample, comprising 4,982 women with at least one live birth in the five years preceding the 2022 GDHS, of whom a majority were rural residents. Most women were also aged 25 to 34 years, with secondary education as the most common level of educational attainment. Nearly a third of women had no formal education, with a pronounced urban-rural disparity: 39.3% of rural women had no education compared with 16.2% of urban women. Women in the analytic sample were predominantly Christian or Muslim. Most women were in a union, working, and with a higher parity. Health insurance coverage was high at 94.7%. Wealth distribution was skewed toward the lower quintiles, and three-quarters of households had electricity, with substantial urban-rural differences (94.6% in urban areas compared with 61.6% in rural areas).

**Table 1.**
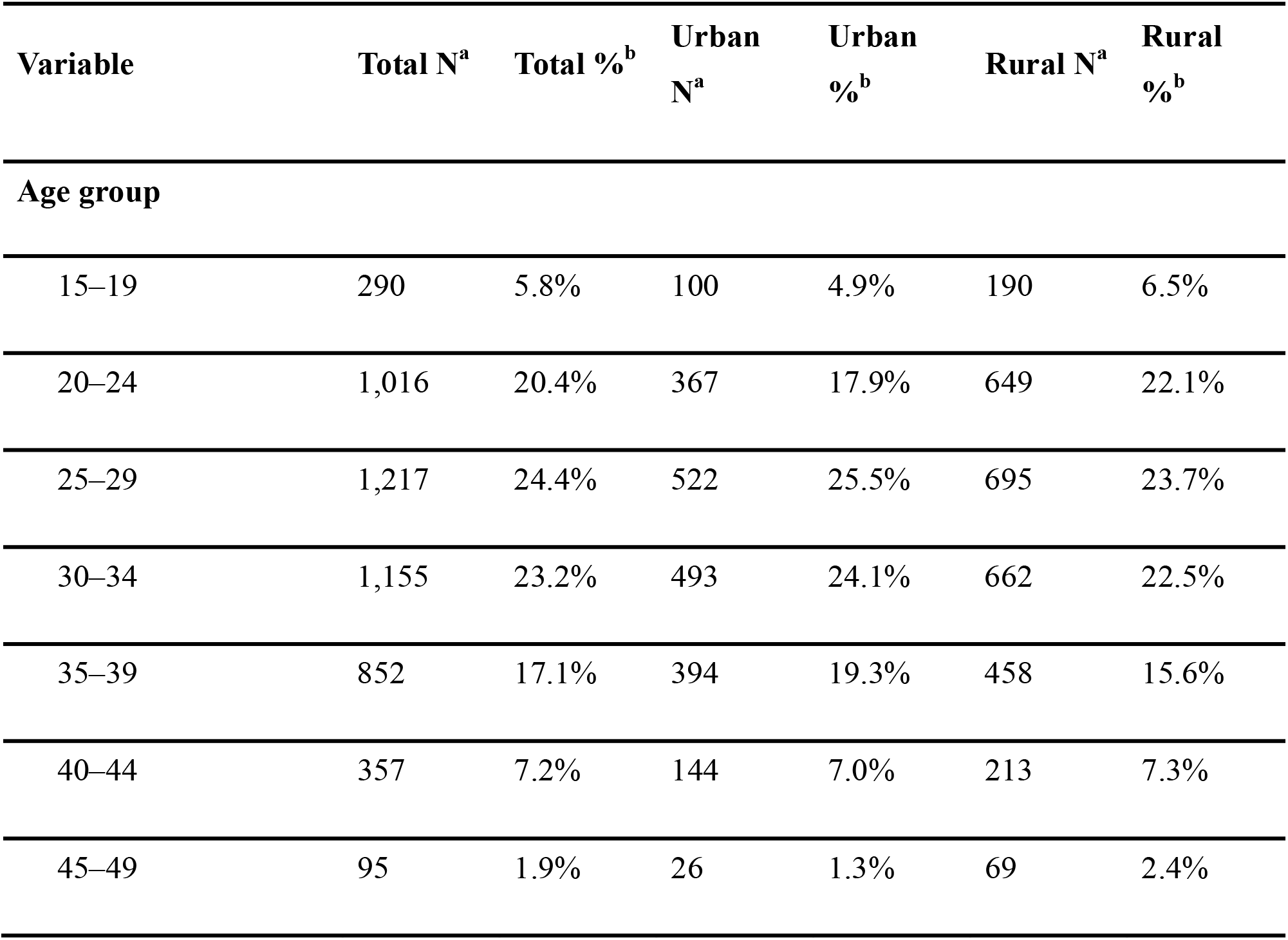
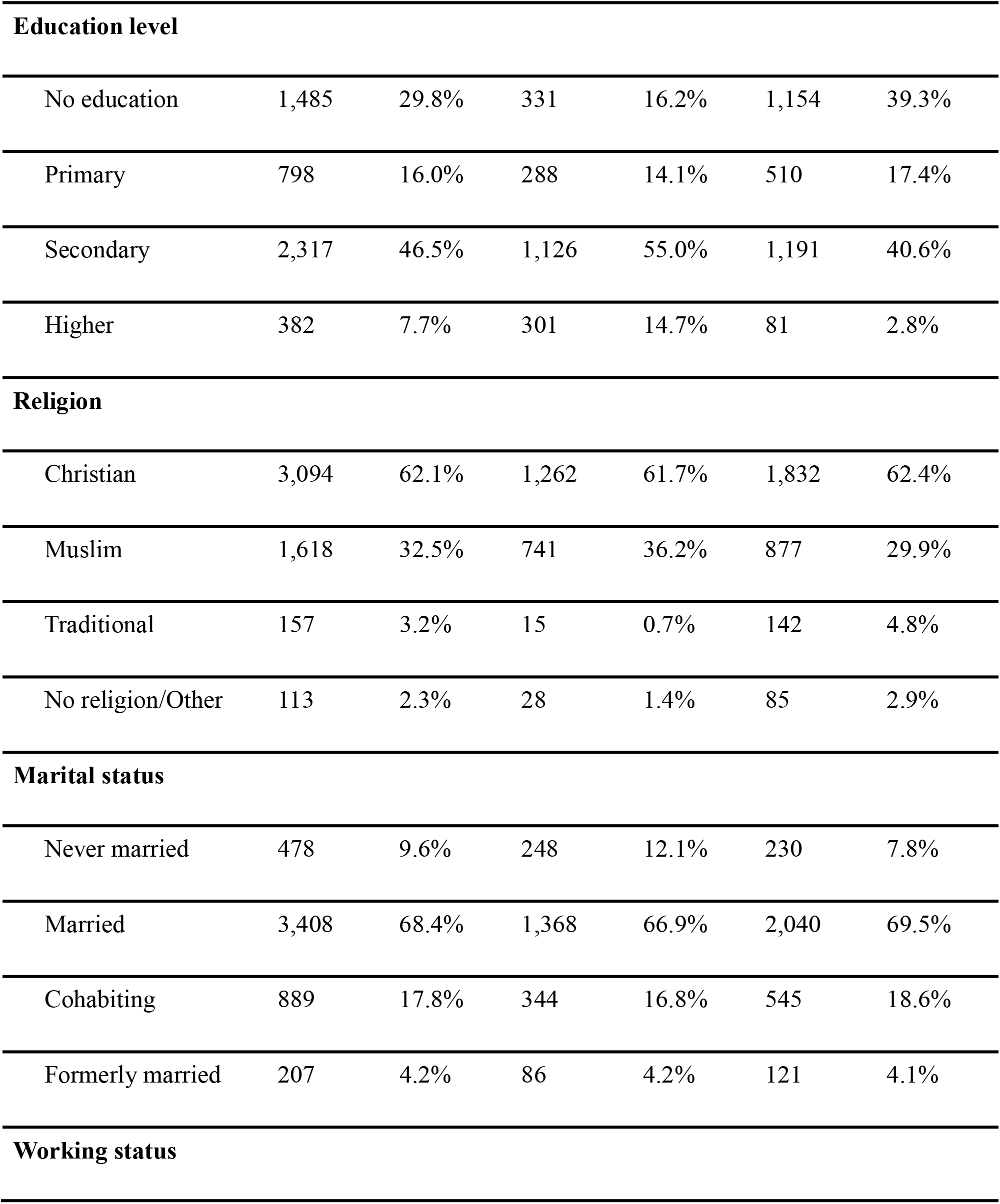
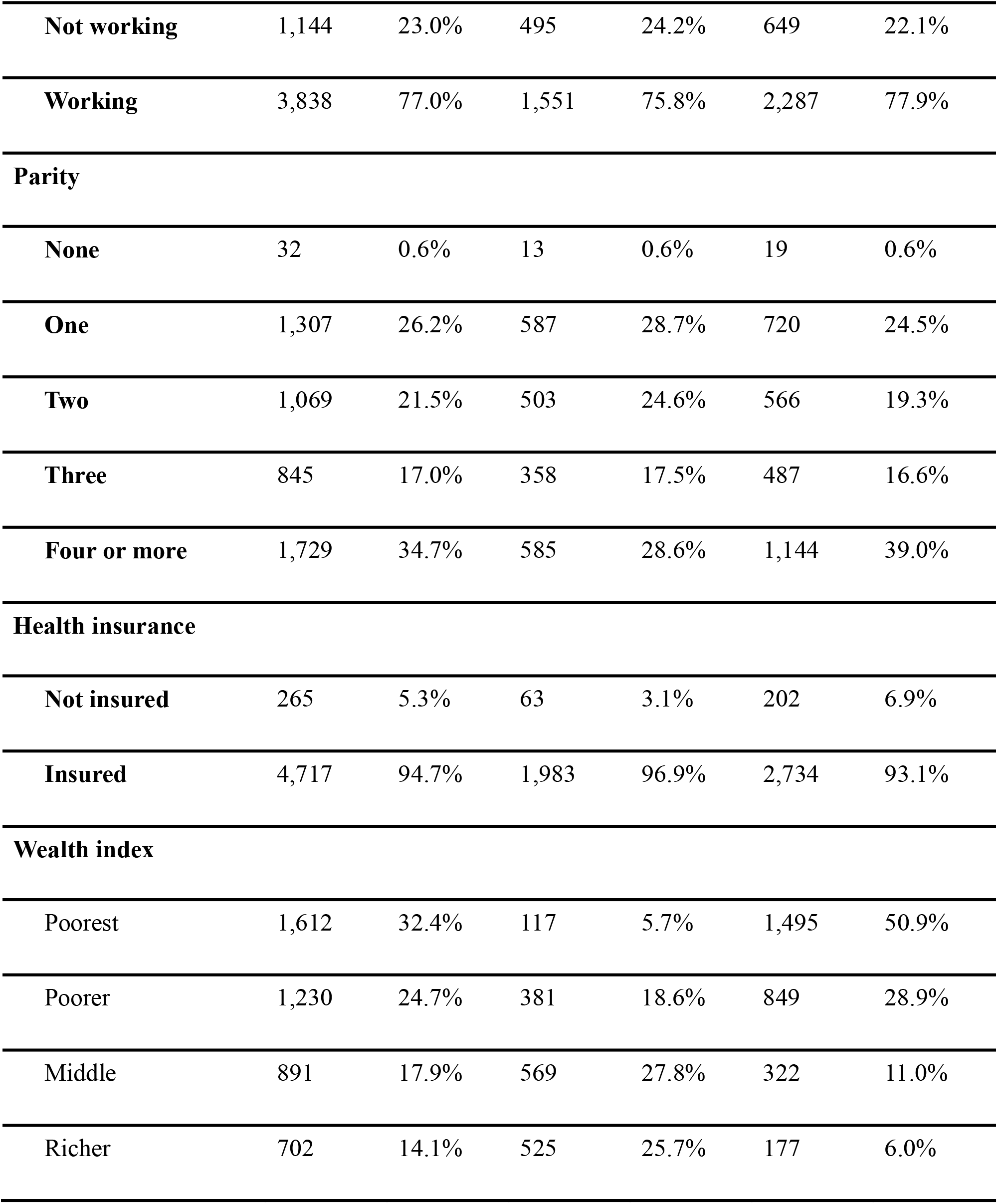
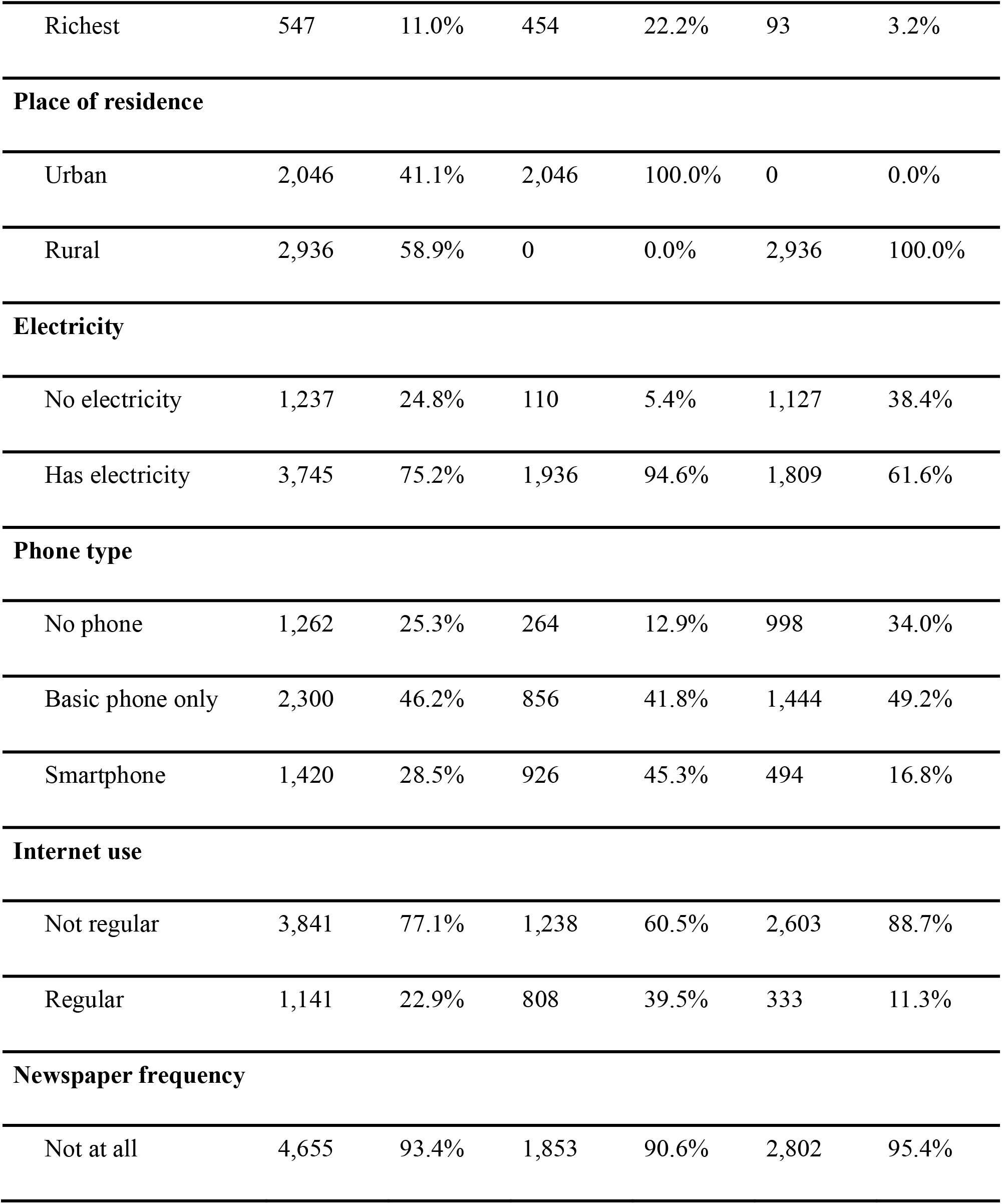
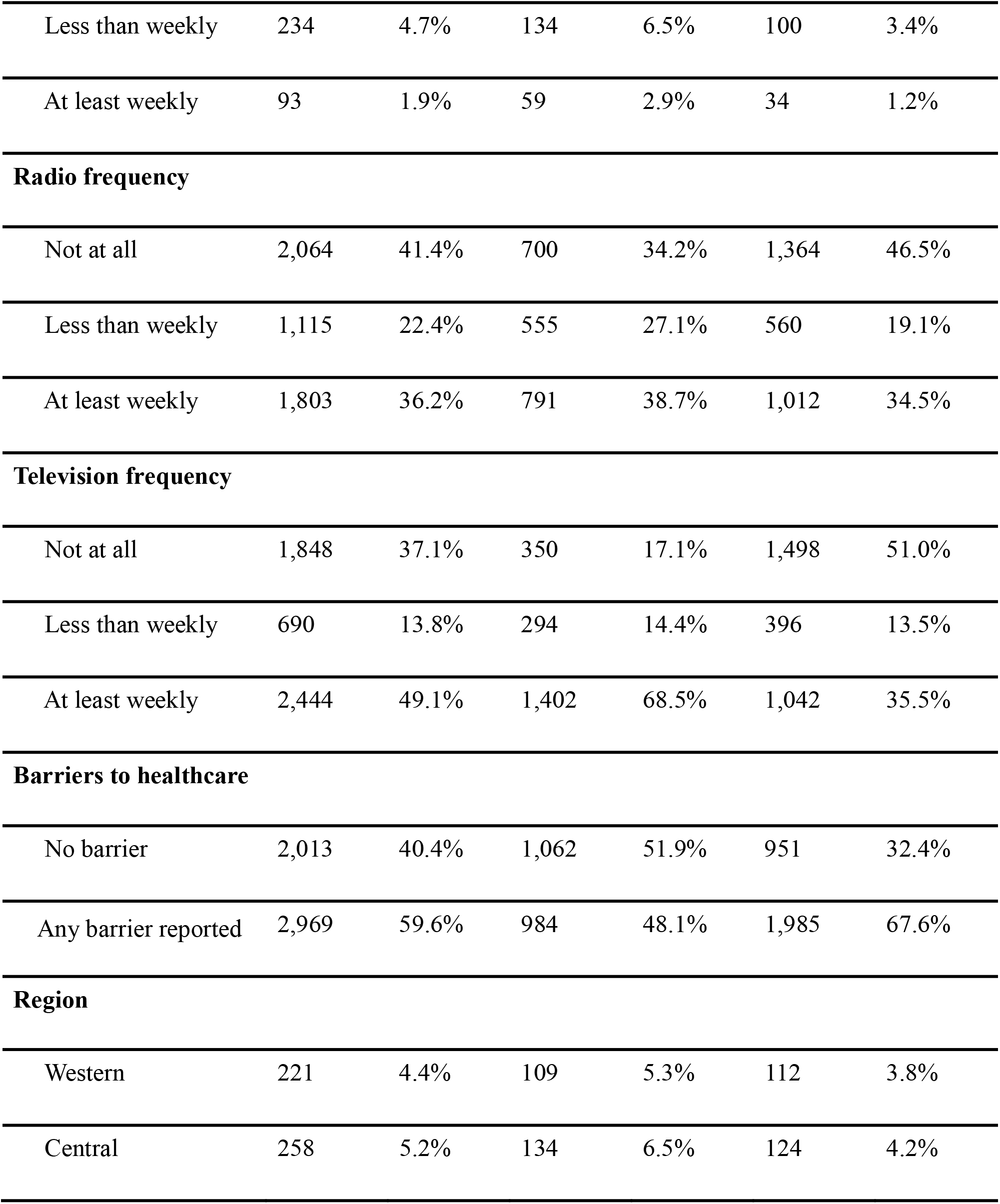
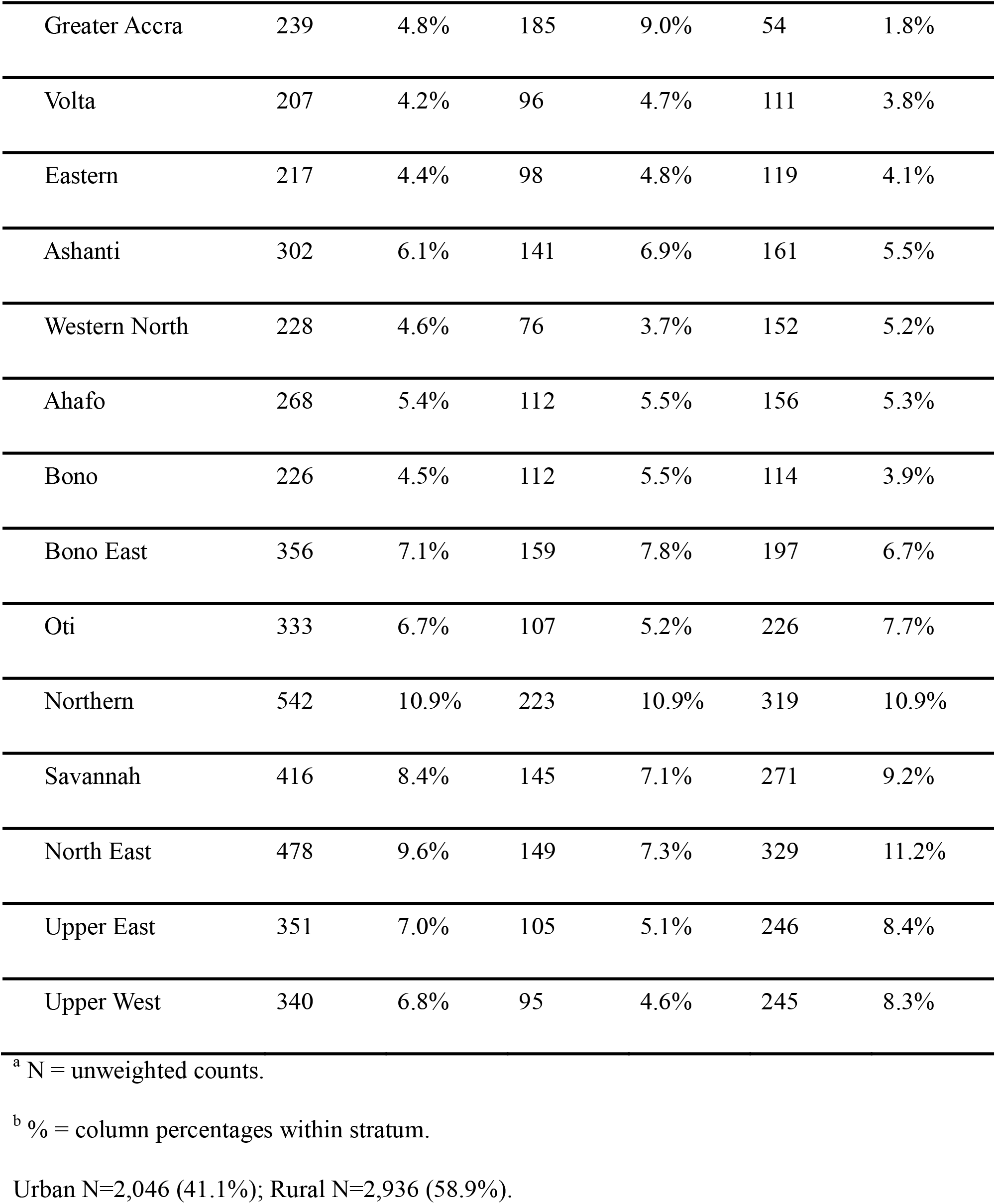
Sociodemographic and reproductive characteristics of women, by place of residence.

| Variable | Total N<sup>a</sup> | Total %<sup>b</sup> | Urban N<sup>a</sup> | Urban %<sup>b</sup> | Rural N<sup>a</sup> | Rural %<sup>b</sup> |
| --- | --- | --- | --- | --- | --- | --- |
| **Age group** |  |  |  |  |  |  |
| 15–19 | 290 | 5.8% | 100 | 4.9% | 190 | 6.5% |
| 20–24 | 1,016 | 20.4% | 367 | 17.9% | 649 | 22.1% |
| 25–29 | 1,217 | 24.4% | 522 | 25.5% | 695 | 23.7% |
| 30–34 | 1,155 | 23.2% | 493 | 24.1% | 662 | 22.5% |
| 35–39 | 852 | 17.1% | 394 | 19.3% | 458 | 15.6% |
| 40–44 | 357 | 7.2% | 144 | 7.0% | 213 | 7.3% |
| 45–49 | 95 | 1.9% | 26 | 1.3% | 69 | 2.4% |

| **Variable** | **Total N<sup>a</sup>** | **Total %<sup>b</sup>** | **Urban**<br>**N<sup>a</sup>** | **Urban**<br>**%<sup>b</sup>** | **Rural N<sup>a</sup>** | **Rural**<br>**%<sup>b</sup>** |
| --- | --- | --- | --- | --- | --- | --- |
| **Education level** |  |  |  |  |  |  |
| No education | 1,485 | 29.8% | 331 | 16.2% | 1,154 | 39.3% |
| Primary | 798 | 16.0% | 288 | 14.1% | 510 | 17.4% |
| Secondary | 2,317 | 46.5% | 1,126 | 55.0% | 1,191 | 40.6% |
| Higher | 382 | 7.7% | 301 | 14.7% | 81 | 2.8% |
| **Religion** |  |  |  |  |  |  |
| Christian | 3,094 | 62.1% | 1,262 | 61.7% | 1,832 | 62.4% |
| Muslim | 1,618 | 32.5% | 741 | 36.2% | 877 | 29.9% |
| Traditional | 157 | 3.2% | 15 | 0.7% | 142 | 4.8% |
| No religion/Other | 113 | 2.3% | 28 | 1.4% | 85 | 2.9% |
| **Marital status** |  |  |  |  |  |  |
| Never married | 478 | 9.6% | 248 | 12.1% | 230 | 7.8% |
| Married | 3,408 | 68.4% | 1,368 | 66.9% | 2,040 | 69.5% |
| Cohabiting | 889 | 17.8% | 344 | 16.8% | 545 | 18.6% |
| Formerly married | 207 | 4.2% | 86 | 4.2% | 121 | 4.1% |
| **Working status** |  |  |  |  |  |  |
| **Not working** | 1,144 | 23.0% | 495 | 24.2% | 649 | 22.1% |
| **Working** | 3,838 | 77.0% | 1,551 | 75.8% | 2,287 | 77.9% |
| **Parity** |  |  |  |  |  |  |
| **None** | 32 | 0.6% | 13 | 0.6% | 19 | 0.6% |
| **One** | 1,307 | 26.2% | 587 | 28.7% | 720 | 24.5% |
| **Two** | 1,069 | 21.5% | 503 | 24.6% | 566 | 19.3% |
| **Three** | 845 | 17.0% | 358 | 17.5% | 487 | 16.6% |
| **Four or more** | 1,729 | 34.7% | 585 | 28.6% | 1,144 | 39.0% |
| **Health insurance** |  |  |  |  |  |  |
| **Not insured** | 265 | 5.3% | 63 | 3.1% | 202 | 6.9% |
| **Insured** | 4,717 | 94.7% | 1,983 | 96.9% | 2,734 | 93.1% |
| **Wealth index** |  |  |  |  |  |  |
| Poorest | 1,612 | 32.4% | 117 | 5.7% | 1,495 | 50.9% |
| Poorer | 1,230 | 24.7% | 381 | 18.6% | 849 | 28.9% |
| Middle | 891 | 17.9% | 569 | 27.8% | 322 | 11.0% |
| Richer | 702 | 14.1% | 525 | 25.7% | 177 | 6.0% |
| Richest | 547 | 11.0% | 454 | 22.2% | 93 | 3.2% |
| **Place of residence** |  |  |  |  |  |  |
| Urban | 2,046 | 41.1% | 2,046 | 100.0% | 0 | 0.0% |
| Rural | 2,936 | 58.9% | 0 | 0.0% | 2,936 | 100.0% |
| **Electricity** |  |  |  |  |  |  |
| No electricity | 1,237 | 24.8% | 110 | 5.4% | 1,127 | 38.4% |
| Has electricity | 3,745 | 75.2% | 1,936 | 94.6% | 1,809 | 61.6% |
| **Phone type** |  |  |  |  |  |  |
| No phone | 1,262 | 25.3% | 264 | 12.9% | 998 | 34.0% |
| Basic phone only | 2,300 | 46.2% | 856 | 41.8% | 1,444 | 49.2% |
| Smartphone | 1,420 | 28.5% | 926 | 45.3% | 494 | 16.8% |
| **Internet use** |  |  |  |  |  |  |
| Not regular | 3,841 | 77.1% | 1,238 | 60.5% | 2,603 | 88.7% |
| Regular | 1,141 | 22.9% | 808 | 39.5% | 333 | 11.3% |
| **Newspaper frequency** |  |  |  |  |  |  |
| Not at all | 4,655 | 93.4% | 1,853 | 90.6% | 2,802 | 95.4% |
| Less than weekly | 234 | 4.7% | 134 | 6.5% | 100 | 3.4% |
| At least weekly | 93 | 1.9% | 59 | 2.9% | 34 | 1.2% |
| **Radio frequency** |  |  |  |  |  |  |
| Not at all | 2,064 | 41.4% | 700 | 34.2% | 1,364 | 46.5% |
| Less than weekly | 1,115 | 22.4% | 555 | 27.1% | 560 | 19.1% |
| At least weekly | 1,803 | 36.2% | 791 | 38.7% | 1,012 | 34.5% |
| **Television frequency** |  |  |  |  |  |  |
| Not at all | 1,848 | 37.1% | 350 | 17.1% | 1,498 | 51.0% |
| Less than weekly | 690 | 13.8% | 294 | 14.4% | 396 | 13.5% |
| At least weekly | 2,444 | 49.1% | 1,402 | 68.5% | 1,042 | 35.5% |
| **Barriers to healthcare** |  |  |  |  |  |  |
| No barrier | 2,013 | 40.4% | 1,062 | 51.9% | 951 | 32.4% |
| Any barrier reported | 2,969 | 59.6% | 984 | 48.1% | 1,985 | 67.6% |
| **Region** |  |  |  |  |  |  |
| Western | 221 | 4.4% | 109 | 5.3% | 112 | 3.8% |
| Central | 258 | 5.2% | 134 | 6.5% | 124 | 4.2% |
| Greater Accra | 239 | 4.8% | 185 | 9.0% | 54 | 1.8% |
| Volta | 207 | 4.2% | 96 | 4.7% | 111 | 3.8% |
| Eastern | 217 | 4.4% | 98 | 4.8% | 119 | 4.1% |
| Ashanti | 302 | 6.1% | 141 | 6.9% | 161 | 5.5% |
| Western North | 228 | 4.6% | 76 | 3.7% | 152 | 5.2% |
| Ahafo | 268 | 5.4% | 112 | 5.5% | 156 | 5.3% |
| Bono | 226 | 4.5% | 112 | 5.5% | 114 | 3.9% |
| Bono East | 356 | 7.1% | 159 | 7.8% | 197 | 6.7% |
| Oti | 333 | 6.7% | 107 | 5.2% | 226 | 7.7% |
| Northern | 542 | 10.9% | 223 | 10.9% | 319 | 10.9% |
| Savannah | 416 | 8.4% | 145 | 7.1% | 271 | 9.2% |
| North East | 478 | 9.6% | 149 | 7.3% | 329 | 11.2% |
| Upper East | 351 | 7.0% | 105 | 5.1% | 246 | 8.4% |
| Upper West | 340 | 6.8% | 95 | 4.6% | 245 | 8.3% |

<sup>a</sup> N = unweighted counts.
<sup>b</sup> % = column percentages within stratum.
Urban N=2,046 (41.1%); Rural N=2,936 (58.9%).

Regarding digital access, approximately one quarter of women owned no mobile phone, and both smartphone ownership and regular internet use were considerably higher among urban women than rural women. Among traditional media, radio and television reached approximately one third and one half of women weekly, respectively. Newspaper readership was low, with only 1.9% reading at least weekly. More than half of women reported at least one barrier to accessing healthcare, with the proportion higher in rural than urban areas.

### Prevalence of maternal health service utilization among women in Ghana

The weighted prevalence of ANC use (four or more visits), adequate ANC (eight or more visits), SBA, and PNC within six weeks among women in Ghana was 88.5%, 38.7%, 86.7%, and 86.3%, respectively (Fig. 1). Urban women had consistently higher utilization across all four outcomes than rural women “S4 Table”. ANC4+ and SBA coverage were both highest in the Upper East Region and lowest in the Oti Region. PNC coverage was also highest in the Upper East Region but lowest in the Northern Region. Adequate ANC coverage followed a different pattern, with the highest coverage recorded in the Western Region and the lowest in the Savannah Region (Fig. 2).

**Figure 1.**
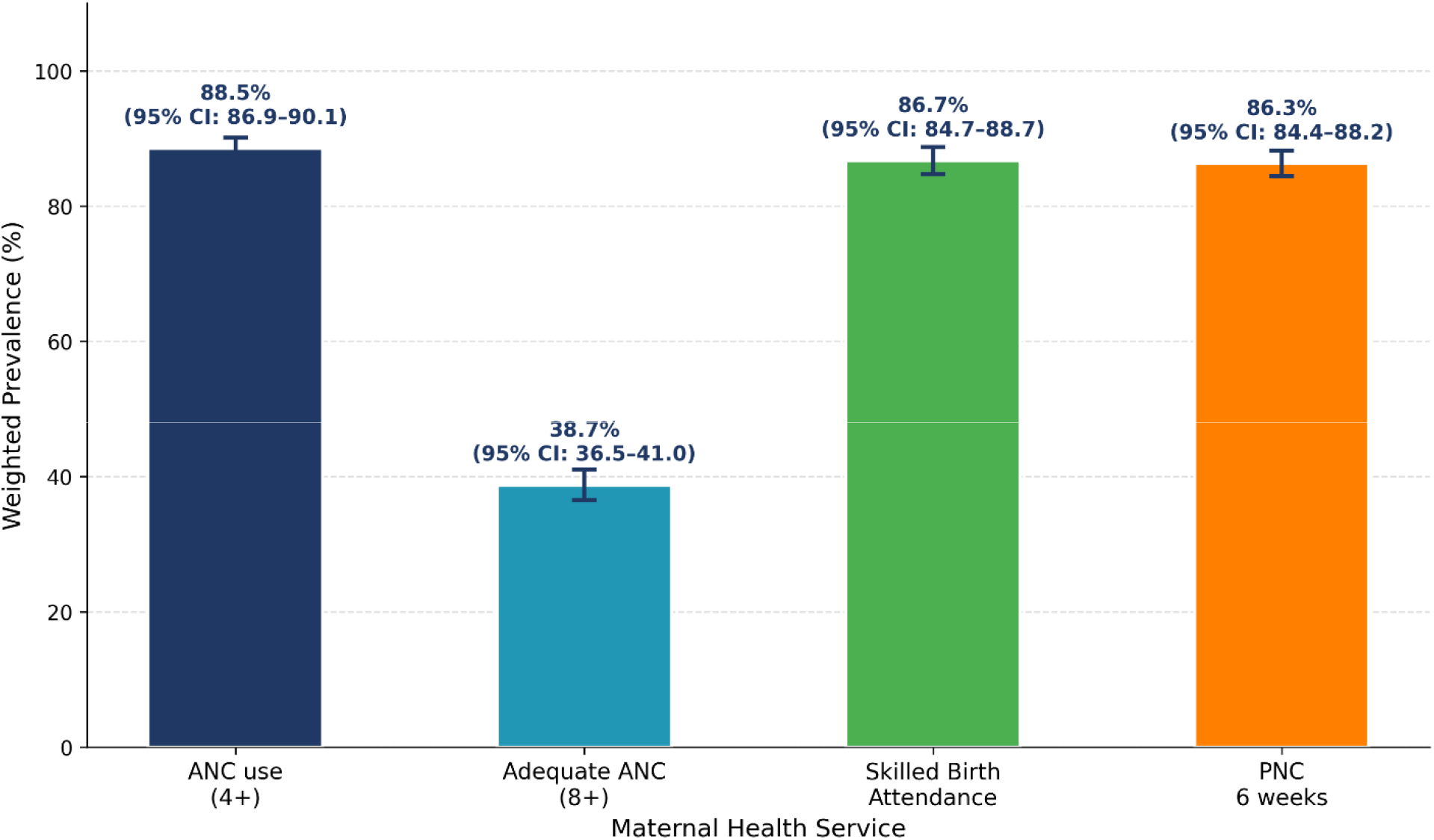
Weighted prevalence of maternal health service utilization among women with a live birth in the five years preceding the 2022 Ghana Demographic and Health Survey. Error bars represent 95% confidence intervals estimated using the complex survey design. ANC use = at least four antenatal care visits; Adequate ANC = at least eight antenatal care visits; SBA = skilled birth attendance by a doctor or nurse/midwife; PNC = postnatal care from a skilled provider within six weeks of delivery.

**Figure 2.**
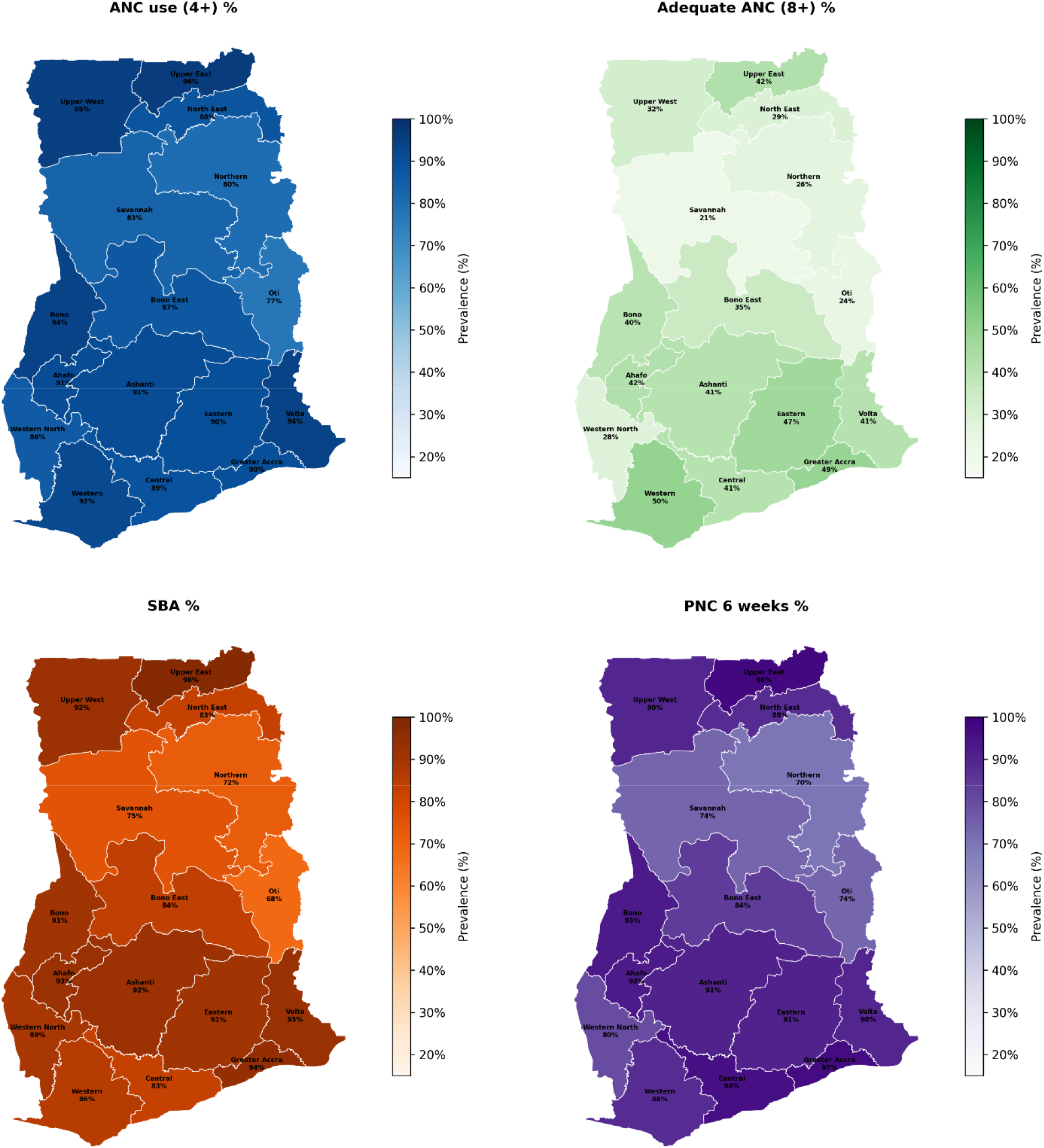
Regional distribution of maternal health service utilization among women with a live birth in the five years preceding the 2022 Ghana Demographic and Health Survey. Maps show survey-weighted prevalences by administrative region. ANC use = at least four antenatal care visits; Adequate ANC = at least eight antenatal care visits; SBA = skilled birth attendance by a doctor or nurse/midwife; PNC = postnatal care from a skilled provider within six weeks of delivery. Regional boundaries sourced from the Global Administrative Areas database (GADM version 4.1)

### Association between explanatory variables and maternal health service utilization

Digital access, media exposure, educational level, religion, health insurance, wealth index, electricity access, place of residence, barriers to healthcare, and region were significantly associated with all four outcomes, with P < 0.001 for each association. Newspaper readership was significantly associated with ANC use (P = 0.003), adequate ANC (P < 0.001), SBA (P < 0.001), and PNC (P = 0.012). No significant associations were observed for age group with SBA (P = 0.096), marital status with adequate ANC (P = 0.171) or PNC (P = 0.090), and working status with SBA (P = 0.086) or PNC (P = 0.472) “S4 Table”.

### Media exposure and maternal health service utilization

After full covariate adjustment, weekly radio listening was the only significant media exposure variable in the total sample, with higher odds of adequate ANC. No media exposure variable was significantly associated with ANC use, SBA, or PNC. Among urban women, no media variable was statistically significant after adjustment. Among rural women, watching television less than once a week showed weak evidence of an association with higher odds of ANC use “Tables 2 and 3”.

**Table 2.**
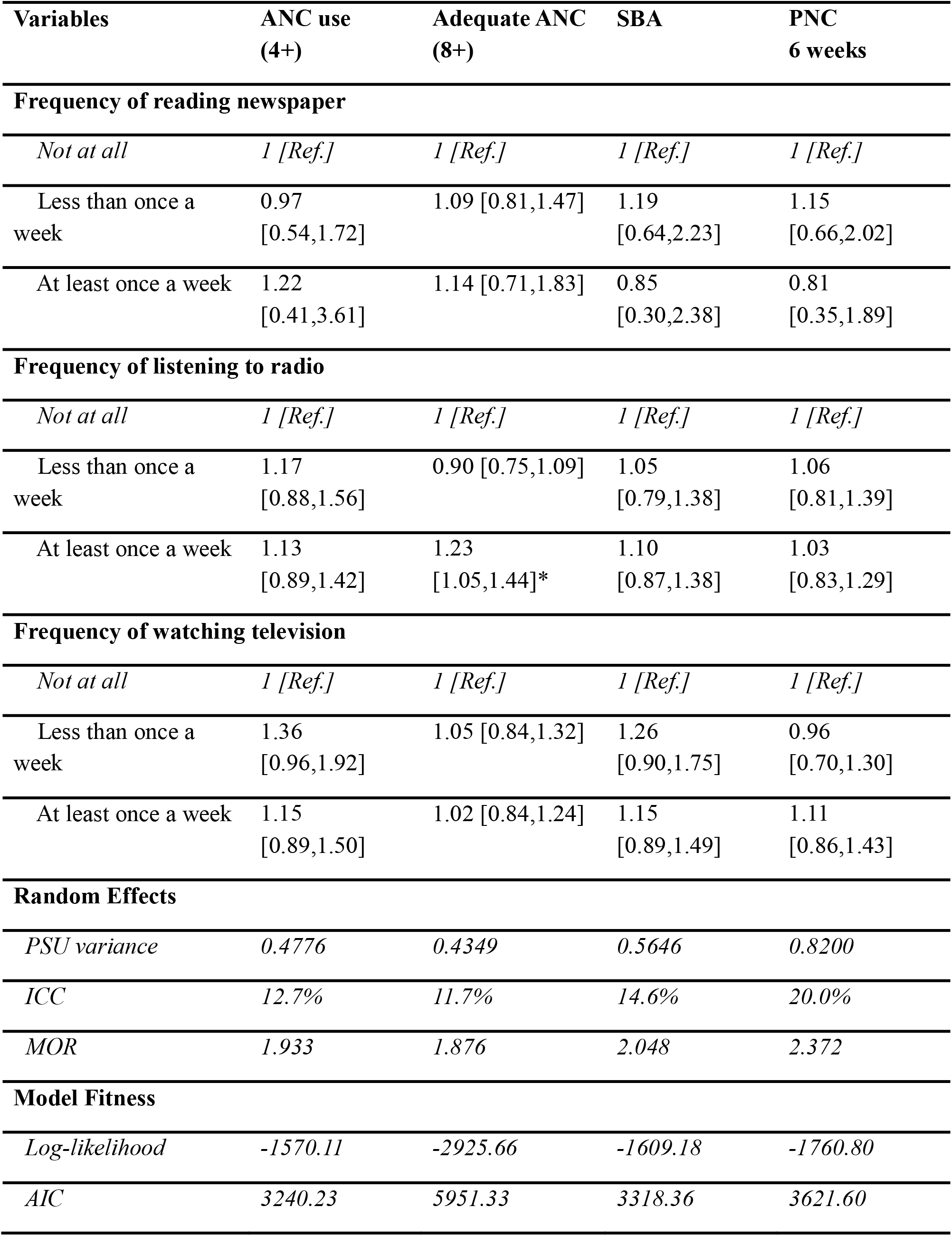
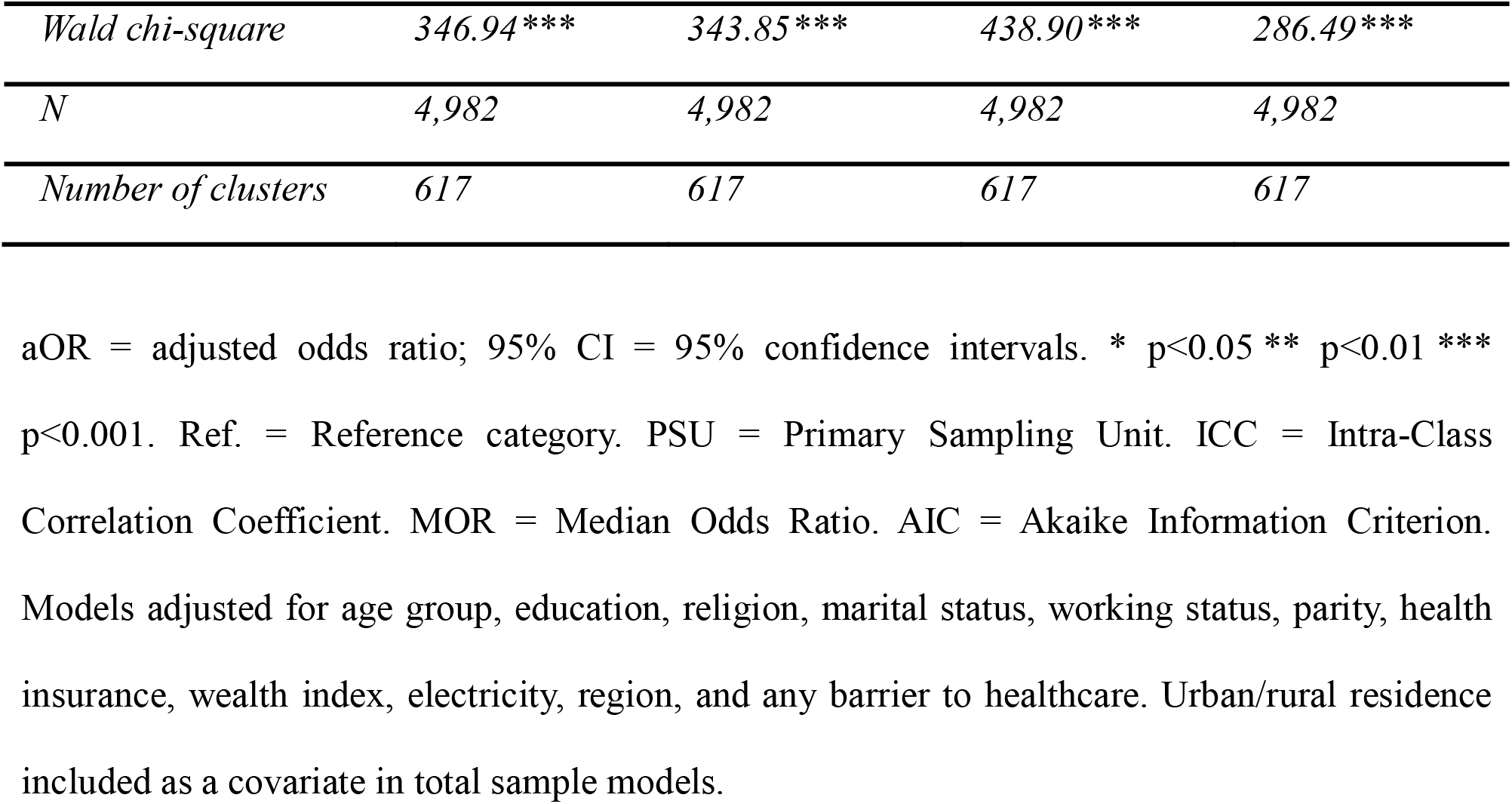
Fixed and random effects results of the association between media exposure and maternal health service utilization among women in Ghana.

**Table 3.**
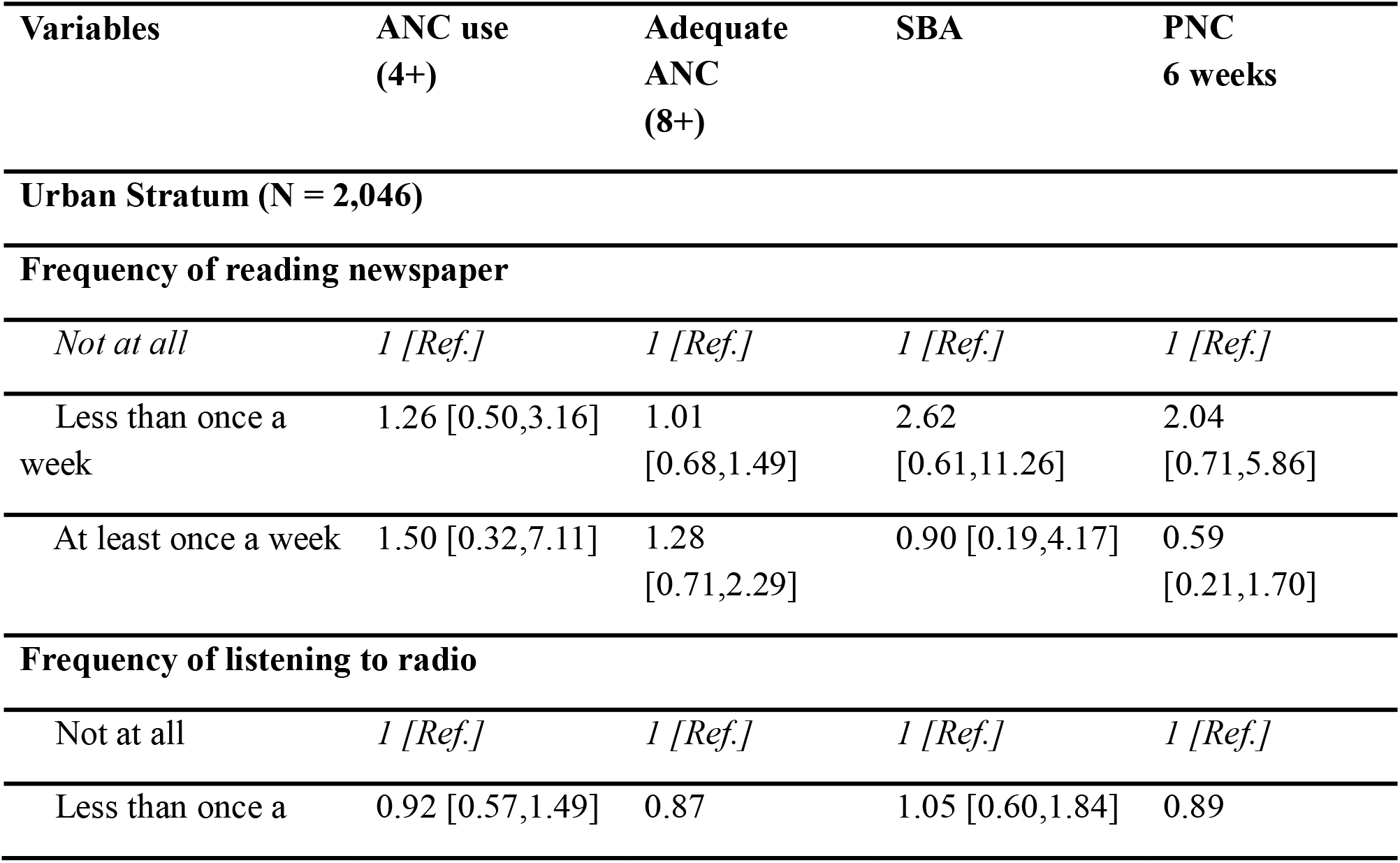
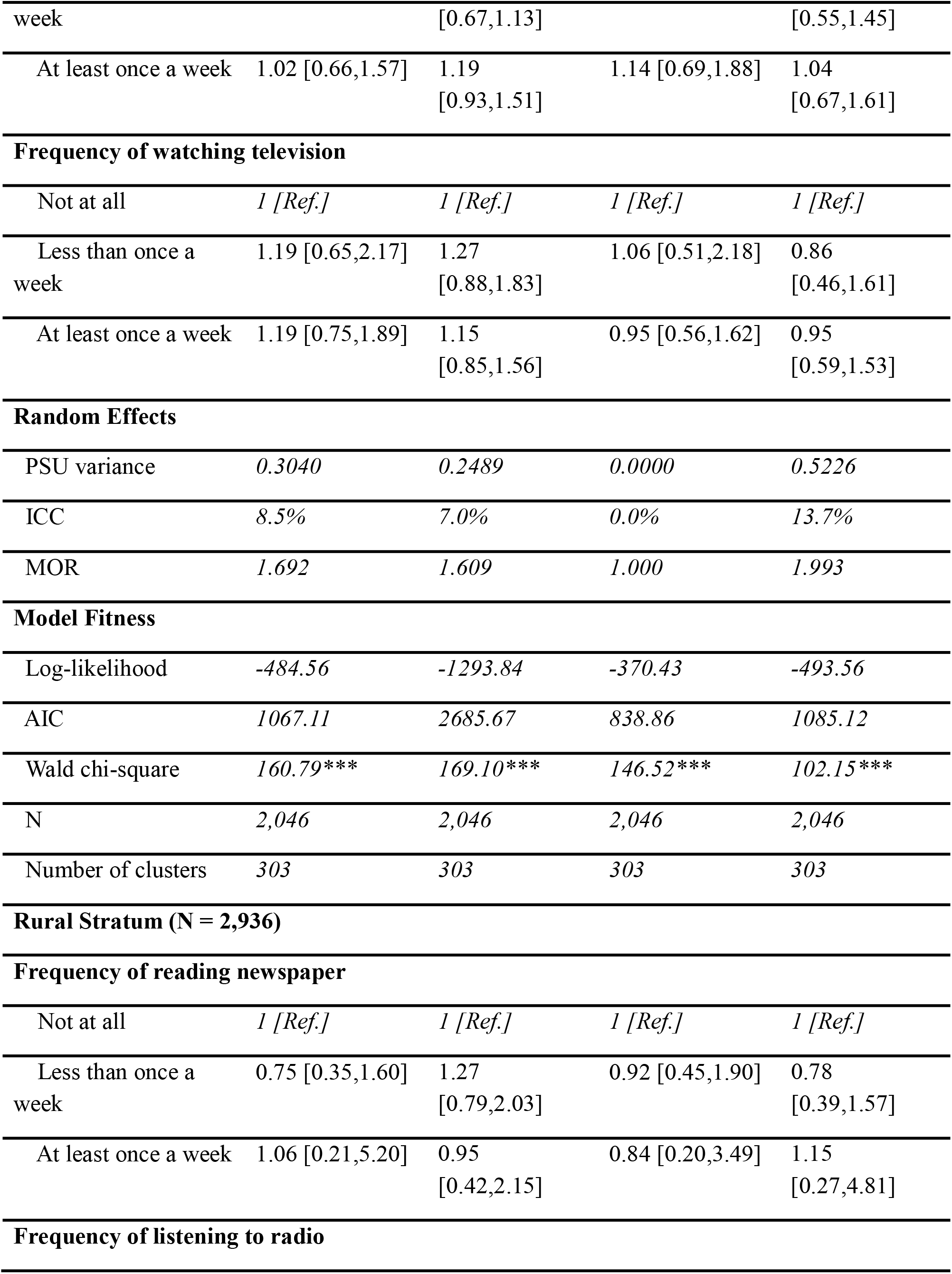
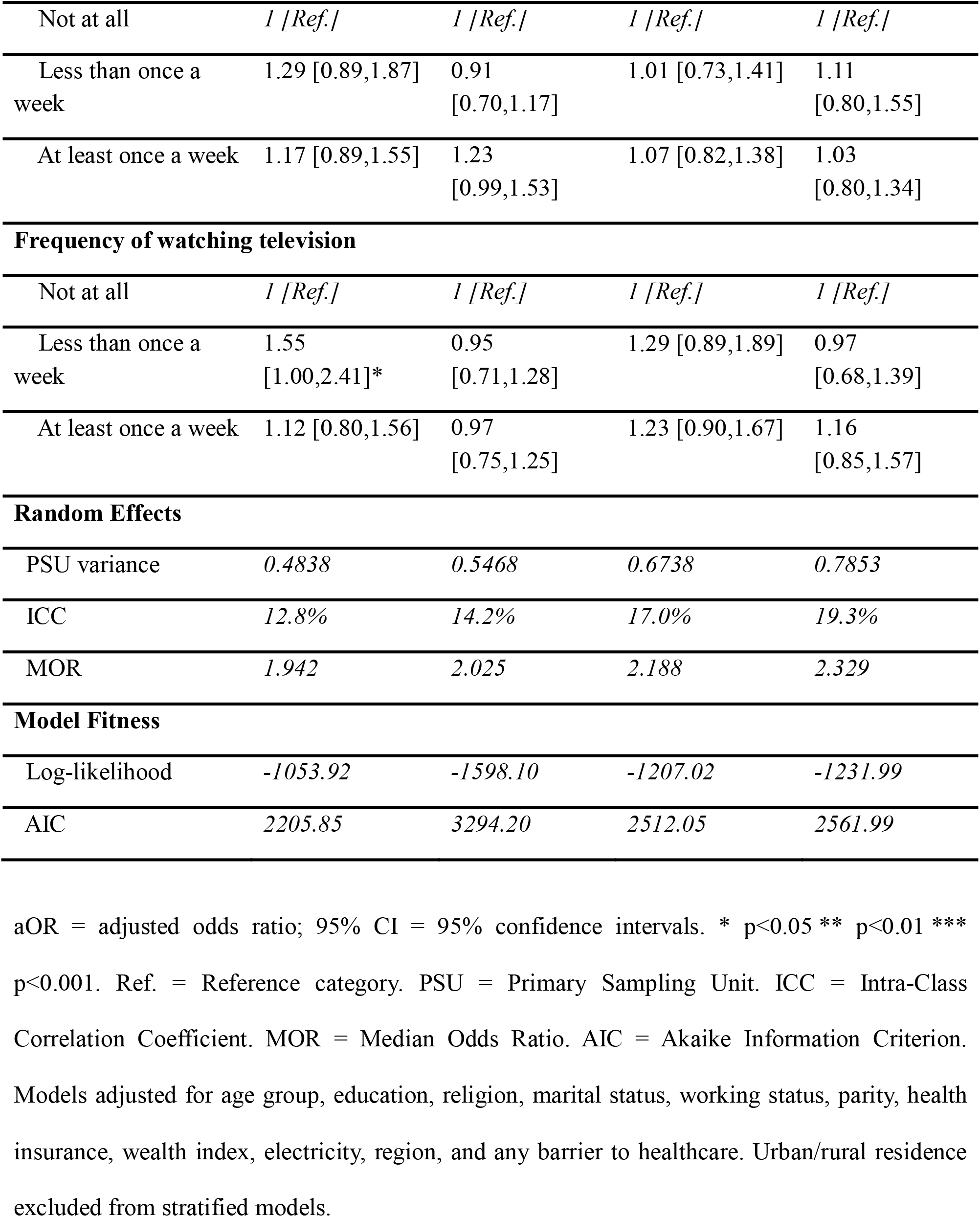
Fixed and random effects results of the association between media exposure and maternal health service utilization - urban and rural strata.

### Digital access and maternal health Service utilization

In the total sample, basic phone ownership was significantly associated with higher odds of ANC use, SBA, and PNC compared with no phone ownership. Smartphone ownership was significantly associated with higher odds of ANC use and SBA. Regular internet use was not independently associated with any outcome. In the urban stratum, smartphone ownership was significantly associated with higher odds of ANC use and SBA; basic phone ownership was significant only for SBA. In the rural stratum, basic phone ownership was significantly associated with higher odds of ANC use, SBA, and PNC; smartphone ownership and internet use were not statistically significant “Tables 4 and 5”.

**Table 4.**
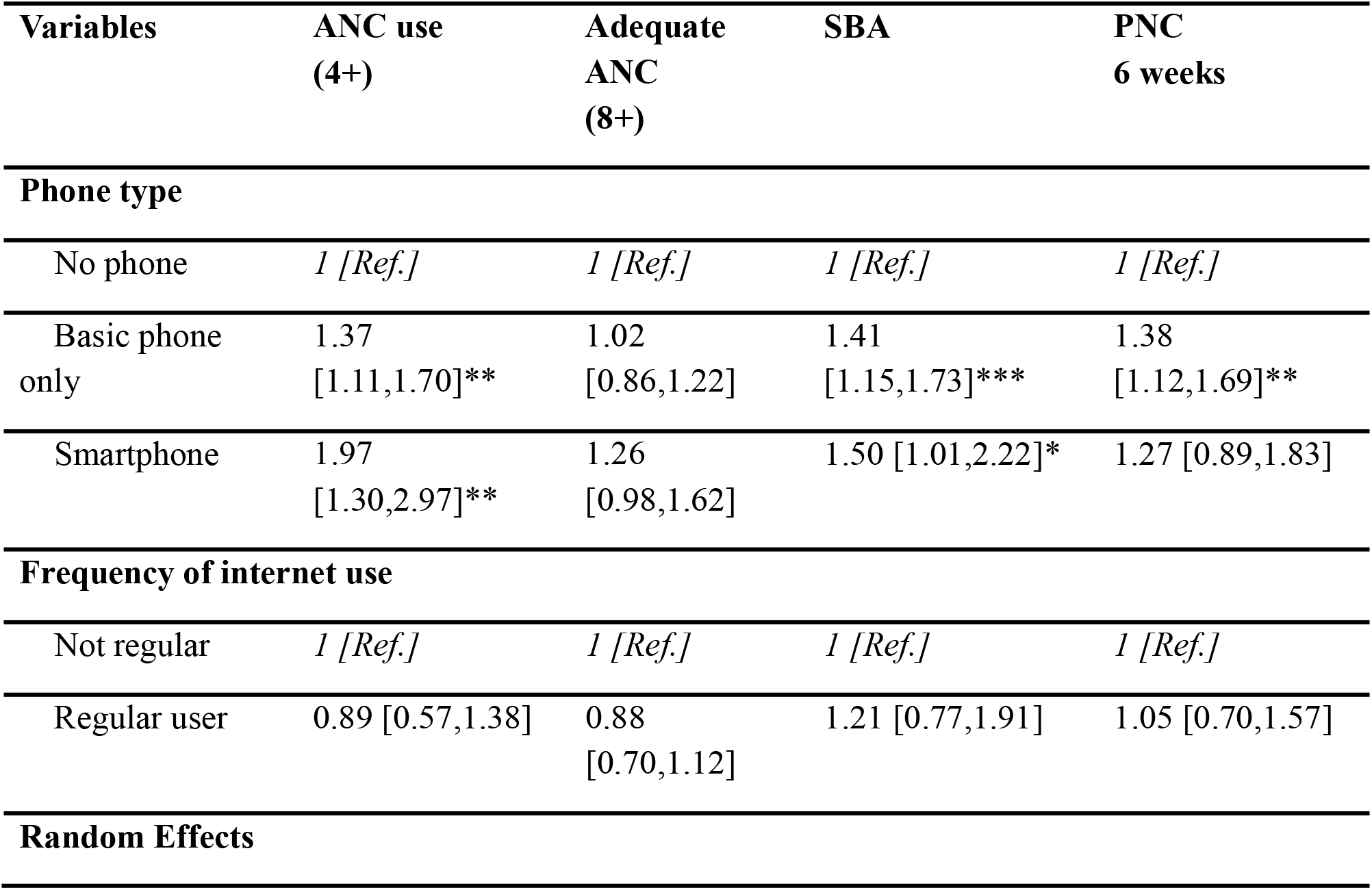
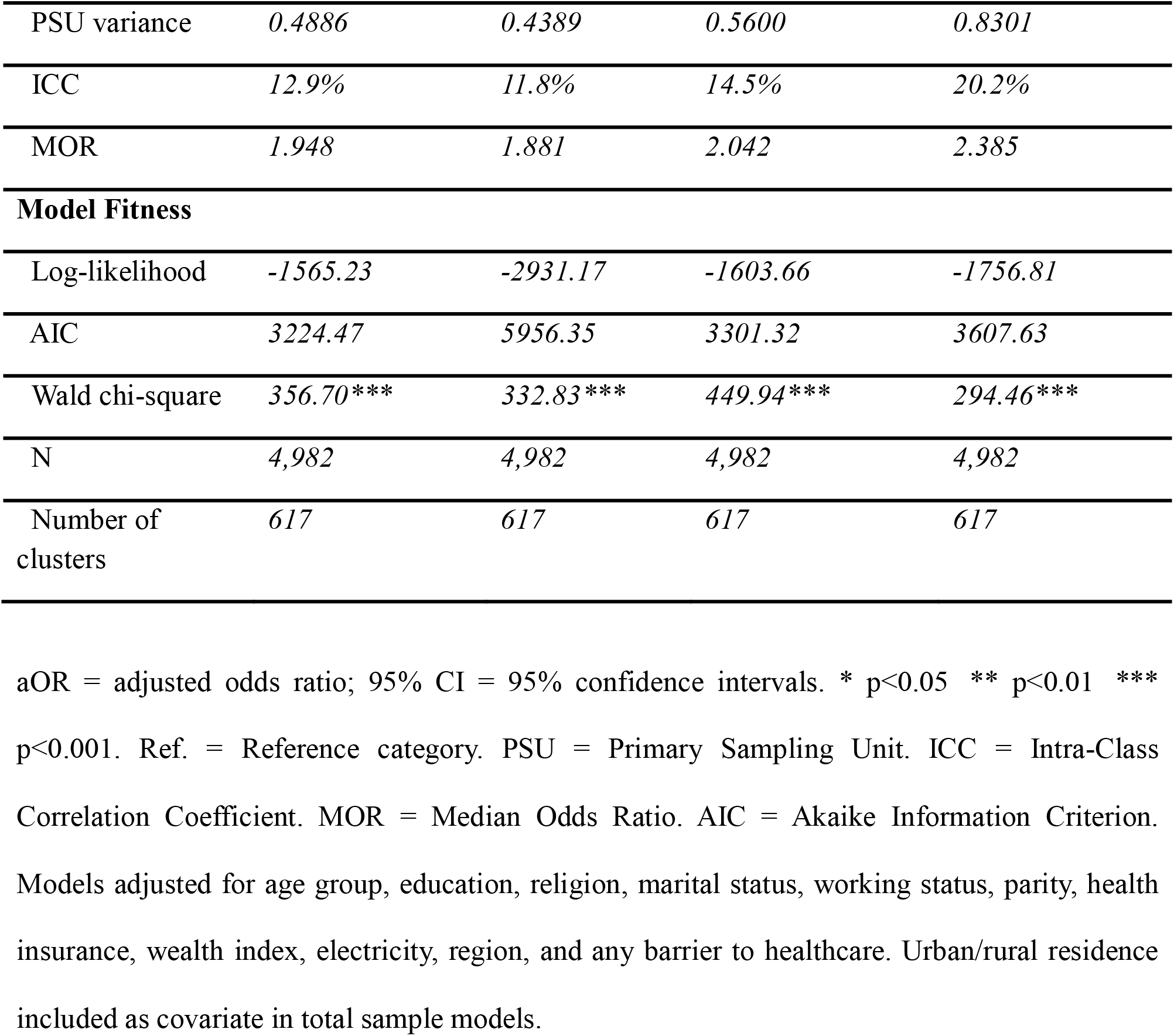
Fixed and random effects results of the association between digital access and maternal health service utilization - total sample.

**Table 5.**
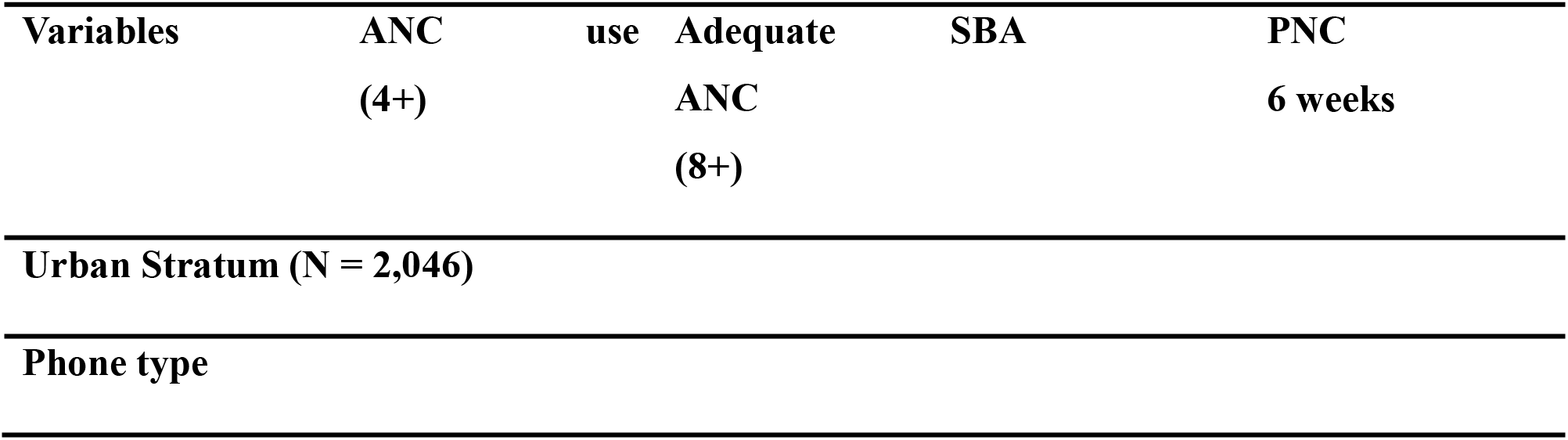
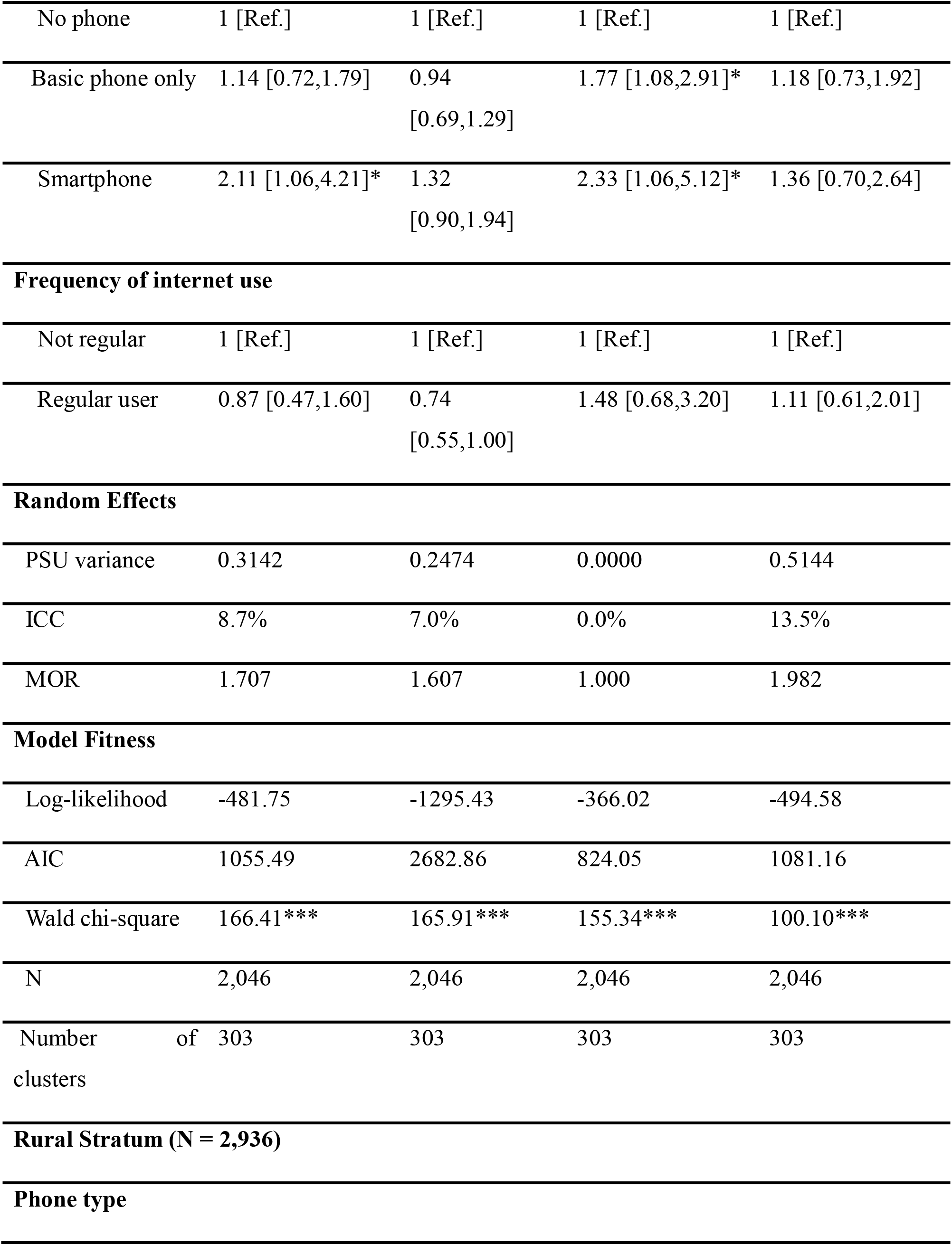
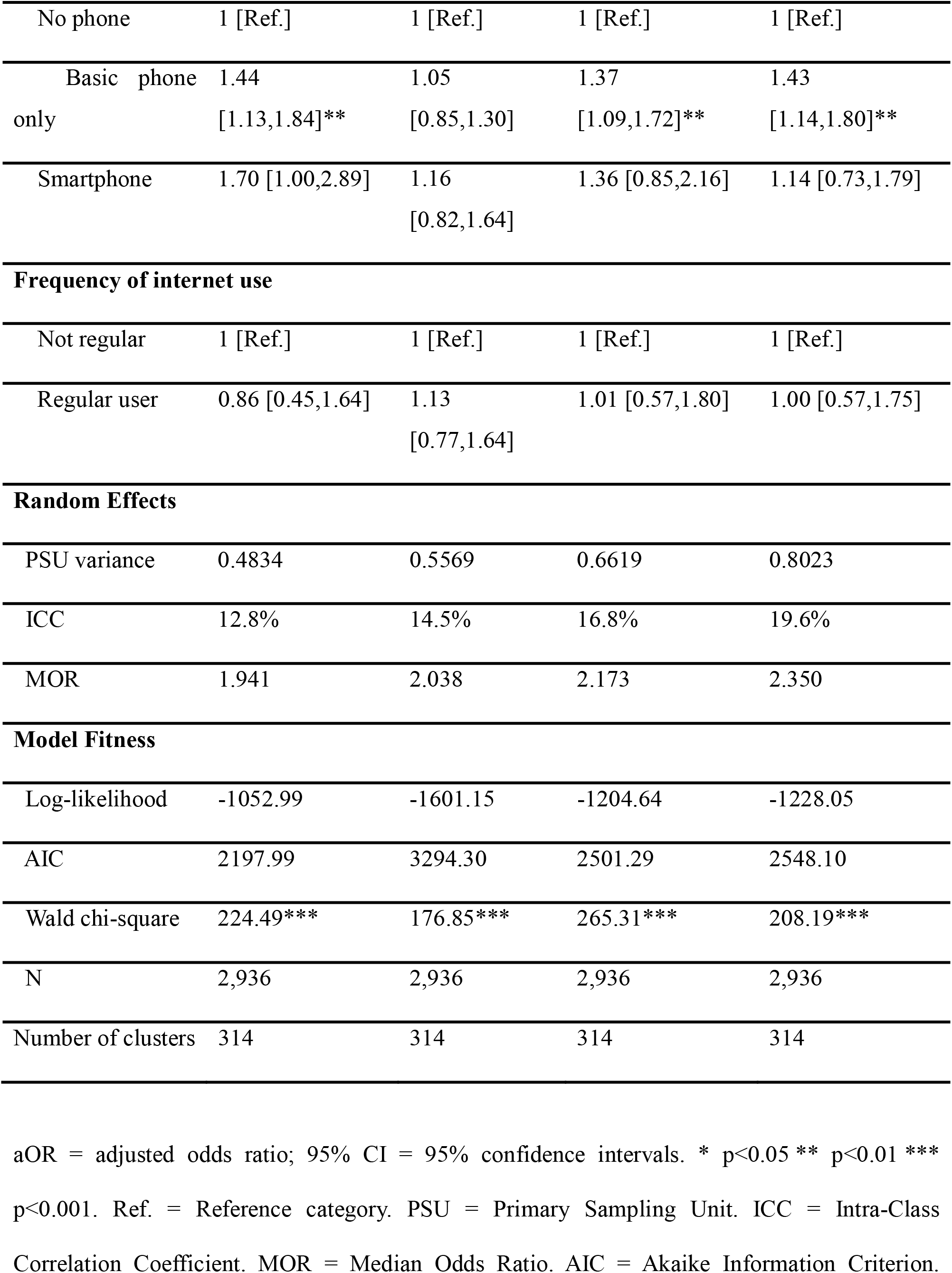
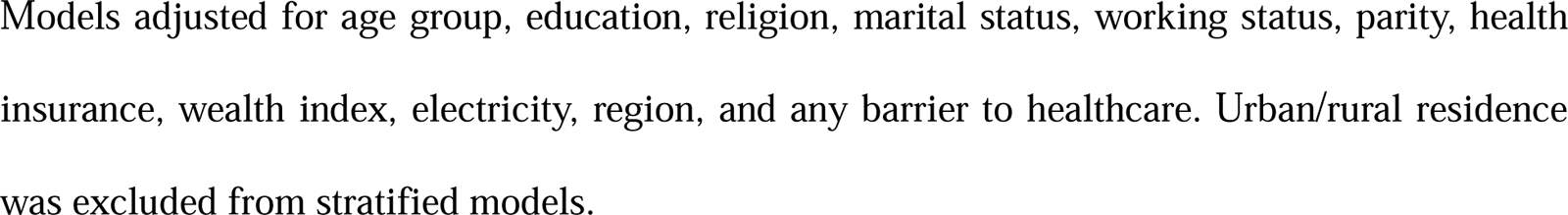
Fixed and random effects results of the association between digital access and maternal health service utilization - urban and rural strata.

## Discussion

This study found that Ghana has achieved high coverage of ANC4+, SBA, and PNC, but substantial urban-rural disparities persist, alongside a national shortfall in achieving the World Health Organization (WHO) recommendation of eight or more antenatal contacts. Urban women were significantly more likely than rural women to achieve ANC4+, deliver with an SBA, and receive PNC, consistent with prior studies in Ghana indicating that inadequate maternal healthcare utilization is concentrated in rural settings [29–31]. The persistent rural disadvantage in maternal healthcare utilization may reflect the concentration of socioeconomic and access-related disadvantages among rural women, who had lower wealth, educational attainment, electricity and digital access, and more healthcare-access barriers “Table 1”. Higher ICCs and MORs in the rural strata “Tables 3 and 5”, further indicate greater residual between-community variation, suggesting that unmeasured contextual factors may also contribute, including transport constraints, healthcare facility accessibility, and workforce availability, as suggested by previous studies [32,33].

In line with the observed disparities in maternal healthcare utilization, the findings also revealed a significant urban-rural digital divide, with urban women considerably more likely than rural women to own smartphones and use the internet regularly. This finding aligns with evidence from Ghana and other low- and middle-income countries showing clear urban-rural disparities in digital access [34,35]. Nevertheless, mobile phone ownership emerged as the most consistent correlate of maternal healthcare utilization across the continuum of care. After adjustment for wealth, education, health insurance, and other socioeconomic characteristics, women who owned basic mobile phones remained significantly more likely to achieve ANC4+, deliver with an SBA, and receive postnatal care than women without mobile phones, while smartphone ownership remained significantly associated with ANC4+ and SBA. These findings are reflective of multi-country evidence from Sub-Saharan Africa demonstrating that mobile phone ownership is associated with greater maternal healthcare engagement [36] and with recent analyses across 39 LMICs showing that mobile phone ownership independently predicts ANC utilization [[37].

Importantly, the relationship between mobile phone ownership and maternal healthcare utilization differed according to both phone type and place of residence. Among rural women, basic mobile phone ownership showed the strongest associations with maternal healthcare utilization, while smartphone ownership was the dominant correlate among urban women. This pattern suggests that the influence of mobile technology on maternal healthcare engagement may be context-specific rather than uniform. In rural settings, where smartphone ownership and digital literacy remain relatively limited, basic mobile phones may facilitate communication through voice calls and SMS, enabling appointment reminders, communication with healthcare providers, and social support [38–40]. Conversely, urban women may benefit more from smartphones because of higher digital literacy and greater access to internet-enabled health information and communication platforms [41]. Although cross-sectional data cannot confirm these mechanisms, they provide plausible explanations for the observed residential differences and merit further investigation.

Unexpectedly, internet use was not independently associated with any maternal healthcare outcome after adjustment, while traditional mass media exposure demonstrated limited independent associations with maternal healthcare utilization. Only weekly radio listening remained significantly associated with adequate ANC in the overall sample, an association driven primarily by rural women. Our results on internet use differed from those of the only Ghana-specific study reporting a significant association between internet exposure and ANC utilization [42]. However, that study focused exclusively on adolescent and young mothers aged 15 to 24 years, limiting direct comparisons with the present study. Several factors may explain the absence of an independent association between internet use, media exposure, and maternal healthcare utilization in our study. First, traditional mass media may be declining in prominence as health communication channels in Ghana, as health professionals and the public have shifted toward digital and mobile platforms for health information [43,44]. Second, the DHS measure captures frequency of internet use regardless of purpose; given documented barriers to online health information seeking in African countries, including low e-health literacy and limited awareness of health-related digital content, general internet use may not translate into engagement with maternal health resources [45,46]. Third, many maternal health communication initiatives in Ghana continue to rely predominantly on voice calls and SMS rather than internet-dependent platforms or mass media [34,35]. Consequently, our findings suggest that internet access and media exposure alone may be insufficient to improve maternal healthcare utilization without purposeful engagement with health-related resources.

Overall, these findings indicate that mobile phone ownership, particularly when considered alongside phone type and residential context, is more strongly associated with maternal healthcare utilization in Ghana than either internet use or traditional mass media exposure alone. The findings thus have several implications for maternal health policy research in Ghana, as they highlight several areas warranting further investigation. The absence of an independent association between internet use and maternal healthcare utilization despite rising internet penetration in Ghana merits exploration in studies with more granular measures of online health information-seeking behavior and e-health literacy. The differential associations observed by phone type and place of residence suggest that the mechanisms linking mobile phone ownership to maternal healthcare utilization may vary by context, and qualitative and mixed-methods studies could help elucidate these pathways. Additionally, the specific association between weekly radio listening and adequate ANC among rural women warrants further examination, including whether particular radio health programs or content types drive this pattern. Future longitudinal research should also investigate the behavioural and communication pathways through which different mobile technologies influence maternal healthcare engagement and evaluate whether targeted mobile-based interventions can reduce persistent urban-rural inequalities in maternal health service utilization in Ghana.

This study has a number of limitations worth noting. First, the cross-sectional design precludes causal inference, and reverse causality or residual confounding cannot be ruled out. Although the multilevel models showed that differences between communities remained after adjustment, the analysis did not include direct measures of community characteristics, such as healthcare facility accessibility, transport infrastructure, or workforce availability. The specific factors underlying these residual between-community differences therefore could not be determined. Second, reliance on self-reported media and digital access data introduces the possibility of recall and social desirability bias; women may over-report or under-report phone ownership, internet use, or media exposure depending on prevailing social norms, and the media exposure variables studied captured only frequency of use rather than the health relevance of content. Third, the DHS phone ownership and media exposure measures do not capture household power dynamics; in many Sub-Saharan African settings, women’s access to and use of mobile phones and media may be mediated or restricted by a partner, such that reported ownership or exposure may not reflect autonomous engagement. Fourth, substantial missing data on outcome variables (26-27%), driven by the DHS questionnaire structure, may introduce some degree of selection bias despite the robustness confirmed by multiple imputation analyses. Finally, the digital access variables reflected ownership and use at the time of the survey, which may not correspond to the period during which women sought maternal care, introducing potential temporal misalignment between exposure and outcome.

## Data Availability

No data was generated by this study. The following existing data source was used: the 2022 Ghana Demographic and Health Survey (GDHS) dataset, available from the DHS Program repository at https://dhsprogram.com upon registration and approval.

## Acknowledgements

The authors thank the DHS Program (ICF International) and the Ghana Statistical Service for making the 2022 Ghana Demographic and Health Survey data publicly available for this analysis.

## Supporting Information

**S1 Table. Variance Inflation Factors (VIF) for All Model Predictors.** Max VIF = 6.00. Mean VIF = 2.43. Values shown reflect Parity merged to 4 categories (see Methods).

**S2 Table. Missingness on Analytic Variables, Eligible Sample (V208**≥**1, N = 6,965).** Missingness on ANC use, adequate ANC, skilled birth attendance, and postnatal care (26.0-27.2% each) predominantly reflects the DHS questionnaire structure: detailed antenatal, delivery, and postnatal care module items are most consistently recorded for births within approximately two years of the survey, with completeness declining for births further in the past within the 5-year eligibility window. Electricity access had minimal missingness (1.5%). All other covariates were fully observed (0% missing). Given the substantial proportion of missingness driven by the outcome variables themselves, multiple imputation by chained equations (m=30) was conducted as a sensitivity analysis “S3 Table”.

**S3 Table. Sensitivity Analysis - Complete Case vs Multiple Imputation by Chained Equations (MICE, m=30, Random Forest, Rubin’s Rules).** aOR = adjusted odds ratio; 95% CI = 95% confidence intervals. * p<0.05 ** p<0.01 *** p<0.001. NS = not significant. Consistent = YES indicates that the complete case and imputed analyses reached the same conclusion of statistical significance (both significant or both non-significant) at p<0.05. Consistent = “Partial” indicates both estimates were significant but crossed different significance thresholds, or one estimate was borderline. No direction reversals were observed. MICE: m=30 imputed datasets, 5 iterations, random forest imputation, Rubin’s rules pooling. Eligible sample: N=6,965.

**S4 Table. Distribution of Maternal Health Service Utilization Across Explanatory Variables.** ^a^Weighted N and % use DHS sampling weights (V005/1,000,000). Prevalences are survey-weighted percentages with 95% confidence intervals in parentheses. ^b^p-values from Rao-Scott adjusted F test accounting for complex survey design.

**S5 Table. Unadjusted Odds Ratios - Media Exposure and Maternal Health Service Utilization (Model I, Total Sample, N = 4,982).** aOR = adjusted odds ratio; 95% CI = 95% confidence intervals. * p<0.05 ** p<0.01 *** p<0.001. Ref. = Reference category. PSU = Primary Sampling Unit. ICC = Intra-Class Correlation. MOR = Median Odds Ratio. AIC = Akaike Information Criterion. Model I: exposure variables only, no covariates.

**S6 Table. Unadjusted Odds Ratios for Media Exposure - Urban and Rural Strata.** aOR = adjusted odds ratio; 95% CI = 95% confidence intervals. * p<0.05 ** p<0.01 *** p<0.001. Ref. = Reference category. PSU = Primary Sampling Unit. ICC = Intra-Class Correlation. MOR = Median Odds Ratio. AIC = Akaike Information Criterion.

**S7 Table. Unadjusted Odds Ratios - Digital Access and Maternal Health Service Utilization (Model I, Total Sample, N = 4,982).** aOR = adjusted odds ratio; 95% CI = 95% confidence intervals. * p<0.05 ** p<0.01 *** p<0.001. Ref. = Reference category. PSU = Primary Sampling Unit. ICC = Intra-Class Correlation. MOR = Median Odds Ratio. AIC = Akaike Information Criterion. Model I: exposure variables only.

**S8 Table. Unadjusted Odds Ratios for Digital Access - Urban and Rural Strata.** aOR = adjusted odds ratio; 95% CI = 95% confidence intervals. * p<0.05 ** p<0.01 *** p<0.001. Ref. = Reference category. PSU = Primary Sampling Unit. ICC = Intra-Class Correlation. MOR = Median Odds Ratio. AIC = Akaike Information Criterion.

**S9 Table. Fully Adjusted Odds Ratios for All Variables - Media Model II (Total Sample, N = 4,982)**. aOR = adjusted odds ratio; 95% CI = 95% confidence intervals. * p<0.05 ** p<0.01 *** p<0.001. Ref. = Reference category. PSU = Primary Sampling Unit. ICC = Intra-Class Correlation. MOR = Median Odds Ratio. AIC = Akaike Information Criterion. Region entered as a 16-category factor (Western Region as reference) per convention in Ghana DHS multilevel analyses.

**S10 Table. Fully Adjusted Odds Ratios for All Variables - Digital Model II (Total Sample, N = 4,982).** aOR = adjusted odds ratio; 95% CI = 95% confidence intervals. * p<0.05 ** p<0.0 *** p<0.001. Ref. = Reference category. PSU = Primary Sampling Unit. ICC = Intra-Class Correlation. MOR = Median Odds Ratio. AIC = Akaike Information Criterion. Region entered as a 16-category factor (Western Region as reference) per convention in Ghana DHS multilevel analyses.

**S1 File. Approval from DHS Program.**

## References

[1] Lawrence ER, Appiah-Kubi A, Lawrence HR, Lui MY, Owusu-Antwi R, Konney T, et al. “There is no joy in the family anymore”: a mixed-methods study on the experience and impact of maternal mortality on families in Ghana 2021. 10.1186/s12884-022-05006-1.

[2] Adu J, Mulay S, Owusu MF. Reducing maternal and child mortality in rural Ghana. Pan Afr Med J 2021;39:263. 10.11604/PAMJ.2021.39.263.30593.

[3] Inusah AW, Nwuzoh M, Seidu A, Ziblim SD. Trends and inequalities in distance related barriers to healthcare access among women of reproductive age in Ghana from 2003 to 2022. Discover Public Health 2026 23:1 2026;23:767-. 10.1186/S12982-026-02117-4.

[4] WHO. Trends in maternal mortality 2000 to 2023: estimates by WHO, UNICEF, UNFPA, World Bank Group and UNDESA/Population Division 2025. https://www.who.int/publications/i/item/9789240108462 (accessed September 8, 2026).

[5] Adu J, Owusu MF. How do we improve maternal and child health outcomes in Ghana? Int J Health Plann Manage 2023;38:898–903. 10.1002/HPM.3639.

[6] Zhao P, Han X, You L, Zhao Y, Yang L, Liu Y. Maternal health services utilization and maternal mortality in China: a longitudinal study from 2009 to 2016. Springer 2020;20:120922. 10.1186/S12884-020-02900-4.

[7] Ahmed T, Roberton T, Vergeer P, Hansen PM, Peters MA, Ofosu AA, et al. Healthcare utilization and maternal and child mortality during the COVID-19 pandemic in 18 low- and middle-income countries: An interrupted time-series analysis with mathematical modeling of administrative data. PLoS Med 2022;19. 10.1371/JOURNAL.PMED.1004070.

[8] Bhutta ZA, Das JK, Bahl R, Lawn JE, Salam RA, Paul VK, et al. Can available interventions end preventable deaths in mothers, newborn babies, and stillbirths, and at what cost? The Lancet 2014;384:347–70. 10.1016/S0140-6736(14)60792-3.

[9] Zelka MA, Yalew AW, Debelew GT. Effectiveness of a continuum of care in maternal health services on the reduction of maternal and neonatal mortality: Systematic review and meta-analysis. Heliyon 2023;9:e17559. 10.1016/J.HELIYON.2023.E17559.

[10] Baten A, Biswas RK, Kendal E, Bhowmik J. Utilization of maternal healthcare services in low- and middle-income countries: a systematic review and meta-analysis. Systematic Reviews 2025 14:1 2025;14:88-. 10.1186/S13643-025-02832-0.

[11] Bobo FT, Asante A, Woldie M, Dawson A, Hayen A. Evaluating equity across the continuum of care for maternal health services: analysis of national health surveys from 25 sub-Saharan African countries. Int J Equity Health 2023;22:239. 10.1186/S12939-023-02047-6.

[12] Ayele BA, Holliday E, Chojenta C. Determinants of antenatal care service utilisation in sub-Saharan Africa: an analysis of demographic and health surveys data (2015–2022). Archives of Public Health 2025;83. 10.1186/S13690-025-01608-1.

[13] Yang Y, Mendoza MH, Adu J. The Association Between Media Exposure and Health Facility Delivery in Ghana 2025. 10.21203/rs.3.rs-6925215/v1.

[14] Abekah-Nkrumah G, Guerriero M, Purohit P. ICTs and maternal healthcare utilization. Evidence from Ghana. Int J Soc Econ 2014;41:518–41. 10.1108/IJSE-11-2012-0218.

[15] Wakefield MA, Loken B, Hornik RC. Use of mass media campaigns to change health behaviour. Lancet 2010;376:1261. 10.1016/S0140-6736(10)60809-4.

[16] Okano JT, Ponce J, Krönke M, Blower S. Lack of ownership of mobile phones could hinder the rollout of mHealth interventions in Africa. Elife 2022;11:e79615. 10.7554/ELIFE.79615.

[17] Anaba EA, Alangea DO, Addo-Lartey A, Modey EJ, Manu A, Alor SK, et al. Determinants of health facility delivery among young mothers in Ghana; insights from the 2014 Ghana Demographic and Health Survey. BMC Pregnancy Childbirth 2022;22. 10.1186/S12884-022-04985-5.

[18] Aboagye RG, Seidu AA, Ahinkorah BO, Cadri A, Frimpong JB, Hagan JE, et al. Association between frequency of mass media exposure and maternal health care service utilization among women in sub-Saharan Africa: Implications for tailored health communication and education. PLoS One 2022;17. 10.1371/JOURNAL.PONE.0275202.

[19] Cai Y. Digital access and health outcomes: The moderating role of socioeconomic status in health information seeking. Digit Health 2026;12:20552076261427776. 10.1177/20552076261427777.

[20] Aboagye RG, Osborne A, Salihu T, Wongnaah FG, Ahinkorah BO. Regional disparities and socio-demographic factors associated with eight or more antenatal care visits in Ghana. Archives of Public Health 2024 82:1 2024;82:192-. 10.1186/S13690-024-01364-8.

[21] Seidu AA, Okyere J, Budu E, Duah HO, Ahinkorah BO. Inequalities in antenatal care in Ghana, 1998–2014. BMC Pregnancy and Childbirth 2022 22:1 2022;22:478-. 10.1186/S12884-022-04803-Y.

[22] Ahinkorah BO, Seidu AA, Agbaglo E, Adu C, Budu E, Hagan JE, et al. Determinants of antenatal care and skilled birth attendance services utilization among childbearing women in Guinea: evidence from the 2018 Guinea Demographic and Health Survey data. BMC Pregnancy and Childbirth 2021 21:1 2021;21:2-. 10.1186/S12884-020-03489-4.

[23] Appiah F, Salihu T, Fenteng JOD, Darteh AO, Kannor P, Ayerakwah PA, et al. Postnatal care utilisation among women in rural Ghana: analysis of 2014 Ghana demographic and health survey. BMC Pregnancy Childbirth 2021;21:26. 10.1186/S12884-020-03497-4.

[24] Edmeades J, MacQuarrie K, Rosenberg R. The relationship between digital access and use and health outcomes: evidence from demographic and health surveys. 2022. 10.5555/20220433111.

[25] Dhawan D, Pinnamaneni R, Bekalu M, Viswanath K. Association between different types of mass media and antenatal care visits in India: a cross-sectional study from the National Family Health Survey (2015–2016). BMJ Open 2020;10:e042839. 10.1136/BMJOPEN-2020-042839.

[26] Fatema K, Lariscy JT. Mass media exposure and maternal healthcare utilization in South Asia. SSM Popul Health 2020;11. 10.1016/J.SSMPH.2020.100614.

[27] Andersen R. A behavioral model of families’ use of health services. 1968. 10.5555/19702701913.

[28] Aheto JMK, Gates T, Tetteh I, Babah R. A multilevel analysis of the predictors of health facility delivery in Ghana: Evidence from the 2014 Demographic and Health Survey. PLOS Global Public Health 2024;4:e0001254. 10.1371/JOURNAL.PGPH.0001254.

[29] Afful-Mensah G, Nketiah-Amponsah E, Boakye-Yiadom L, Afful-Mensah G, Nketiah-Amponsah E, Boakye-Yiadom L. Rural-Urban Differences in the Utilization of Maternal Healthcare in Ghana: The Case of Antenatal and Delivery Services. African Social Science Review 2014;6:4.

[30] Nuamah GB, Agyei-Baffour P, Mensah KA, Boateng D, Quansah DY, Dobin D, et al. Access and utilization of maternal healthcare in a rural district in the forest belt of Ghana. Springer 2019;19. 10.1186/S12884-018-2159-5.

[31] Anarwat SG, Salifu M, Akuriba MA. Equity and access to maternal and child health services in Ghana a cross-sectional study. Springer 2021;21. 10.1186/S12913-021-06872-9.

[32] Bassoumah B, Adam AM, Adokiya MN. Challenges to the utilization of Community-based Health Planning and Services: the views of stakeholders in Yendi Municipality, Ghana. BMC Health Serv Res 2021;21:1223. 10.1186/S12913-021-07249-8.

[33] Ganle KK, Parker M, Fitzpatrick R, Otupiri E. A qualitative study of health system barriers to accessibility and utilization of maternal and newborn healthcare services in Ghana after user-fee abolition. BMC Pregnancy and Childbirth 2014 14:1 2014;14:425-. 10.1186/S12884-014-0425-8.

[34] Oduro-Mensah E, Agyepong IA, Frimpong E, Zweekhorst M, Vanotoo LA. Implementation of a referral and expert advice call Center for Maternal and Newborn Care in the resource constrained health system context of the Greater Accra region of Ghana. BMC Pregnancy and Childbirth 2021 21:1 2021;21:56-. 10.1186/S12884-020-03534-2.

[35] Willcox M, Moorthy A, Mohan D, Romano K, Hutchful D, Mehl G, et al. Mobile Technology for Community Health in Ghana: Is Maternal Messaging and Provider Use of Technology Cost-Effective in Improving Maternal and Child Health Outcomes at Scale? J Med Internet Res 2019;21:e11268. 10.2196/11268.

[36] Iacoella F, Gassmann F, Tirivayi N. Which communication technology is effective for promoting reproductive health? Television, radio, and mobile phones in sub-Saharan Africa. PLoS One 2022;17:e0272501. 10.1371/JOURNAL.PONE.0272501.

[37] Rafi A-D, Syeda Zobia N, Sameen Z, Muhammad K, Rao Muhammad A. Women’s mobile connectivity: improving ANC care utilisation in developing countries. Discover Health Systems 2025 4:1 2025;4:113-. 10.1007/S44250-025-00230-W.

[38] Peprah P, Abalo EM, Agyemang-Duah W, Budu HI, Appiah-Brempong E, Morgan AK, et al. Lessening barriers to healthcare in rural Ghana: providers and users’ perspectives on the role of mHealth technology. A qualitative exploration. BMC Med Inform Decis Mak 2020;20:27. 10.1186/S12911-020-1040-4.

[39] Omole O, Ijadunola MY, Olotu E, Omotoso O, Bello B, Awoniran O, et al. The effect of mobile phone short message service on maternal health in south-west Nigeria. International Journal of Health Planning and Management 2018;33:155–70. 10.1002/HPM.2404

[40] De P, Pradhan MR. Effectiveness of mobile technology and utilization of maternal and neonatal healthcare in low and middle-income countries (LMICs): a systematic review. Springer 2023;23. 10.1186/S12905-023-02825-Y.

[41] Qin C, Zhu Y, Li D, Liu C. The impact of digital skills on health: Evidence from the China General Social Survey. Digit Health 2024;10. 10.1177/20552076241304592

[42] Anaba EA, Alor SK, Badzi CD. Utilization of antenatal care among adolescent and young mothers in Ghana; analysis of the 2017/2018 multiple indicator cluster survey. BMC Pregnancy Childbirth 2022;22. 10.1186/S12884-022-04872-Z.

[43] Adekunle TB, Mohammed WF. Communication in context: How culture, structure, and agency shape health and risk communication about COVID 19 in Ghana. World Med Health Policy 2022;14:437. 10.1002/WMH3.522.

[44] Bannor R, Asare AK, Bawole JN. Effectiveness of social media for communicating health messages in Ghana. Health Educ 2017;117:342–71. 10.1108/HE-06-2016-0024.

[45] GSMA. Understanding Mobile Internet Use in Low- and Middle-Income Countries – The State of Mobile Internet Connectivity 2025 2025.

[46] Chereka AA, Shibabaw AA, Butta FW, Tadesse MN, Abebe MT, Atanie FA, et al. Explore barriers to using the internet for health information access in African countries: A systematic review. PLOS Digital Health 2025;4:e0000719. 10.1371/JOURNAL.PDIG.0000719.

