## Supplementary file for "Digital access, media exposure, and maternal health service utilization among women in Ghana"

May 06, 2026

Sandy Baffour Adu
Summer Health Limited
Ghana
Request Date: 05/06/2026

Dear Sandy Baffour Adu:

This is to confirm that you are approved to use the following Survey Datasets for your registered research project titled: "Digital Access, Media Exposure, and Maternal Health Service Utilization in Ghana: An Analysis of the 2022 Ghana Demographic and Health Survey (GDHS)":

**Ghana**

To access the datasets, please login at: https://www.dhsprogram.com/data/dataset_admin/login_main.cfm. The user name is the registered email address, and the password is the one selected during registration.

The IRB-approved procedures for DHS public-use datasets do not in any way allow respondents, households, or sample communities to be identified. There are no names of individuals or household addresses in the data files. The geographic identifiers only go down to the regional level (where regions are typically very large geographical areas encompassing several states/provinces). Each enumeration area (Primary Sampling Unit) has a PSU number in the data file, but the PSU numbers do not have any labels to indicate their names or locations. In surveys that collect GIS coordinates in the field, the coordinates are only for the enumeration area (EA) as a whole, and not for individual households, and the measured coordinates are randomly displaced within a large geographic area so that specific enumeration areas cannot be identified.

The DHS Data may be used only for the purpose of statistical reporting and analysis, and only for your registered research. To use the data for another purpose, a new research project must be registered. All DHS data should be treated as confidential, and no effort should be made to identify any household or individual respondent interviewed in the survey. Also, be aware that re-distribution of any DHS micro-level data, either directly or within any tool/dashboard, is not permitted. Please reference the complete terms of use at: https://dhsprogram.com/Data/terms-of-use.cfm.

The data must not be passed on to other researchers without the written consent of DHS. However, if you have coresearchers registered in your account for this research paper, you are authorized to share the data with them. All data users are required to submit an electronic copy (pdf) of any reports/publications resulting from using the DHS data files to:.

Sincerely,

Olsen Hanner

Olsen Hanner
Data Archivist
The Demographic and Health Surveys (DHS) Program
