## Supplementary table for "Digital access, media exposure, and maternal health service utilization among women in Ghana"

**S1 Table. Variance Inflation Factors (VIF) for All Model Predictors**

| **Variable** | **VIF** |
| --- | --- |
| **Age_group_4.0** | 6.003 |
| **Age_group_5.0** | 5.460 |
| **Age_group_3.0** | 5.189 |
| **Age_group_2.0** | 3.968 |
| **Region_12.0** | 3.822 |
| **Parity_4** | 3.776 |
| **Wealth_index_5.0** | 3.660 |
| **Marital_status_1** | 3.611 |
| **Region_14.0** | 3.560 |
| **Wealth_index_4.0** | 3.230 |
| **Age_group_6.0** | 3.196 |
| **Phone_type_2** | 3.140 |
| **Wealth_index_3.0** | 3.088 |
| **Region_13.0** | 3.079 |
| **Region_15.0** | 2.706 |
| **Region_16.0** | 2.702 |
| **Marital_status_2** | 2.657 |
| **Region_10.0** | 2.639 |
| **Internet_use** | 2.544 |
| **Region_11.0** | 2.522 |
| **Wealth_index_2.0** | 2.509 |
| **Electricity** | 2.387 |
| **Education_2.0** | 2.301 |
| **Region_6.0** | 2.274 |
| **Region_8.0** | 2.223 |
| **Education_3.0** | 2.137 |
| **Television_2** | 2.137 |
| **Region_2.0** | 2.099 |
| **Region_3.0** | 2.044 |
| **Region_7.0** | 2.007 |
| **Region_9.0** | 1.990 |
| **Parity_3** | 1.984 |
| **Region_5.0** | 1.943 |
| **Region_4.0** | 1.898 |
| **Parity_2** | 1.721 |
| **Urban_rural** | 1.672 |
| **Phone_type_1** | 1.665 |
| **Age_group_7.0** | 1.650 |
| **Religion_2** | 1.534 |
| **Education_1.0** | 1.512 |
| **Marital_status_3** | 1.475 |
| **Television_1** | 1.467 |
| **Radio_2** | 1.400 |
| **Radio_1** | 1.387 |
| **Religion_3** | 1.235 |
| **Any_barrier** | 1.177 |
| **Working_status** | 1.124 |
| **Newspaper_2** | 1.084 |
| **Newspaper_1** | 1.080 |
| **Religion_4** | 1.054 |
| **Health_insurance** | 1.052 |

Max VIF = 6.00. Mean VIF = 2.43. Values shown reflect Parity merged to 4 categories (see Methods).
