## Supplementary table for "Digital access, media exposure, and maternal health service utilization among women in Ghana"

**S2 Table. Missingness on Analytic Variables, Eligible Sample (V208≥1, N = 6,965)**

| **Variable** | **N Missing** | **% Missing** |
| --- | --- | --- |
| **Outcome variables** | | |
| ANC use (4+ visits) | 1,827 | 26.2% |
| Adequate ANC (8+ visits) | 1,827 | 26.2% |
| Skilled birth attendance | 1,814 | 26.0% |
| Postnatal care (6 weeks) | 1,892 | 27.2% |
| **Digital access** | | |
| Phone type | 0 | 0.0% |
| Internet use | 0 | 0.0% |
| **Media exposure** | | |
| Newspaper frequency | 0 | 0.0% |
| Radio frequency | 0 | 0.0% |
| Television frequency | 0 | 0.0% |
| **Sociodemographic covariates** | | |
| Age group | 0 | 0.0% |
| Educational level | 0 | 0.0% |
| Religion | 0 | 0.0% |
| Marital status | 0 | 0.0% |
| Working status | 0 | 0.0% |
| Parity | 0 | 0.0% |
| Health insurance | 0 | 0.0% |
| Wealth index | 0 | 0.0% |
| Place of residence | 0 | 0.0% |
| Electricity access | 106 | 1.5% |
| Region | 0 | 0.0% |
| Any barrier to healthcare | 0 | 0.0% |
| **Overall** | | |
| Missing on ≥1 analytic variable | 1,983 | 28.5% |
| Complete cases (analytic sample) | 4,982 | 71.5% |

Missingness on ANC use, adequate ANC, skilled birth attendance, and postnatal care (26.0-27.2% each) predominantly reflects the DHS questionnaire structure: detailed antenatal, delivery, and postnatal care module items are most consistently recorded for births within approximately two years of the survey, with completeness declining for births further in the past within the 5-year eligibility window. Electricity access had minimal missingness (1.5%). All other covariates were fully observed (0% missing). Given the substantial proportion of missingness driven by the outcome variables themselves, multiple imputation by chained equations (m=30) was conducted as a sensitivity analysis.
