## Supplementary table for "Digital access, media exposure, and maternal health service utilization among women in Ghana"

**S3 Table. Sensitivity Analysis - Complete Case vs Multiple Imputation by Chained Equations (MICE, m=30, Random Forest, Rubin's Rules)**

| **Model** | **Variable** | **Outcome** | **Complete Case aOR [95% CI]** | **Imputed (m=30) aOR [95% CI]** | **Agree?** |
| --- | --- | --- | --- | --- | --- |
| **Media** | | | | | |
|  | Newspaper <weekly | ANC use (4+) | 0.97 [0.54,1.72]NS | 0.93 [0.53,1.61]NS | Yes |
|  | Newspaper ≥weekly | ANC use (4+) | 1.22 [0.41,3.61]NS | 1.28 [0.45,3.67]NS | Yes |
|  | Radio <weekly | ANC use (4+) | 1.17 [0.88,1.56]NS | 1.14 [0.86,1.50]NS | Yes |
|  | Radio ≥weekly | ANC use (4+) | 1.13 [0.89,1.42]NS | 1.17 [0.94,1.46]NS | Yes |
|  | TV <weekly | ANC use (4+) | 1.36 [0.96,1.92]NS | 1.33 [0.97,1.83]NS | Yes |
|  | TV ≥weekly | ANC use (4+) | 1.15 [0.89,1.50]NS | 1.15 [0.90,1.46]NS | Yes |
|  | Newspaper <weekly | Adequate ANC (8+) | 1.09 [0.81,1.47]NS | 1.03 [0.78,1.34]NS | Yes |
|  | Newspaper ≥weekly | Adequate ANC (8+) | 1.14 [0.71,1.83]NS | 1.23 [0.79,1.91]NS | Yes |
|  | Radio <weekly | Adequate ANC (8+) | 0.90 [0.75,1.09]NS | 0.95 [0.80,1.12]NS | Yes |
|  | Radio ≥weekly | Adequate ANC (8+) | 1.23 [1.05,1.44]* | 1.28 [1.10,1.49]** | Partial |
|  | TV <weekly | Adequate ANC (8+) | 1.05 [0.84,1.32]NS | 1.05 [0.84,1.32]NS | Yes |
|  | TV ≥weekly | Adequate ANC (8+) | 1.02 [0.84,1.24]NS | 1.05 [0.88,1.25]NS | Yes |
|  | Newspaper <weekly | SBA | 1.19 [0.64,2.23]NS | 0.97 [0.55,1.71]NS | Yes |
|  | Newspaper ≥weekly | SBA | 0.85 [0.30,2.38]NS | 0.80 [0.31,2.08]NS | Yes |
|  | Radio <weekly | SBA | 1.05 [0.79,1.38]NS | 1.04 [0.81,1.34]NS | Yes |
|  | Radio ≥weekly | SBA | 1.10 [0.87,1.38]NS | 1.13 [0.92,1.40]NS | Yes |
|  | TV <weekly | SBA | 1.26 [0.90,1.75]NS | 1.31 [0.97,1.78]NS | Yes |
|  | TV ≥weekly | SBA | 1.15 [0.89,1.49]NS | 1.24 [0.97,1.58]NS | Yes |
|  | Newspaper <weekly | PNC 6 weeks | 1.15 [0.66,2.02]NS | 1.00 [0.60,1.68]NS | Yes |
|  | Newspaper ≥weekly | PNC 6 weeks | 0.81 [0.35,1.89]NS | 0.84 [0.38,1.85]NS | Yes |
|  | Radio <weekly | PNC 6 weeks | 1.06 [0.81,1.39]NS | 1.02 [0.80,1.30]NS | Yes |
|  | Radio ≥weekly | PNC 6 weeks | 1.03 [0.83,1.29]NS | 1.05 [0.85,1.29]NS | Yes |
|  | TV <weekly | PNC 6 weeks | 0.96 [0.70,1.30]NS | 1.07 [0.81,1.41]NS | Yes |
|  | TV ≥weekly | PNC 6 weeks | 1.11 [0.86,1.43]NS | 1.19 [0.94,1.49]NS | Yes |
| **Digital** | | | | | |
|  | Basic phone | ANC use (4+) | 1.37 [1.11,1.70]** | 1.33 [1.09,1.63]** | Yes |
|  | Smartphone | ANC use (4+) | 1.97 [1.30,2.97]** | 2.01 [1.34,3.02]*** | Partial |
|  | Regular internet | ANC use (4+) | 0.89 [0.57,1.38]NS | 0.91 [0.61,1.37]NS | Yes |
|  | Basic phone | Adequate ANC (8+) | 1.02 [0.86,1.22]NS | 1.01 [0.86,1.19]NS | Yes |
|  | Smartphone | Adequate ANC (8+) | 1.26 [0.98,1.62]NS | 1.27 [1.01,1.59]* | Partial |
|  | Regular internet | Adequate ANC (8+) | 0.88 [0.70,1.12]NS | 0.90 [0.73,1.11]NS | Yes |
|  | Basic phone | SBA | 1.41 [1.15,1.73]*** | 1.41 [1.16,1.71]*** | Yes |
|  | Smartphone | SBA | 1.50 [1.01,2.22]* | 1.70 [1.17,2.47]** | Partial |
|  | Regular internet | SBA | 1.21 [0.77,1.91]NS | 1.02 [0.66,1.57]NS | Yes |
|  | Basic phone | PNC 6 weeks | 1.38 [1.12,1.69]** | 1.33 [1.11,1.61]** | Yes |
|  | Smartphone | PNC 6 weeks | 1.27 [0.89,1.83]NS | 1.34 [0.95,1.88]NS | Yes |
|  | Regular internet | PNC 6 weeks | 1.05 [0.70,1.57]NS | 0.96 [0.66,1.40]NS | Yes |

aOR = adjusted odds ratio; 95% CI = 95% confidence intervals. * p<0.05 ** p<0.01 *** p<0.001. NS = not significant. Consistent = YES indicates that the complete case and imputed analyses reached the same conclusion of statistical significance (both significant or both non-significant) at p<0.05. Consistent = "Partial" indicates both estimates were significant but crossed different significance thresholds, or one estimate was borderline. No direction reversals were observed. MICE: m=30 imputed datasets, 5 iterations, random forest imputation, Rubin's rules pooling. Eligible sample: N=6,965.
