## Supplementary table for "Digital access, media exposure, and maternal health service utilization among women in Ghana"

**S4 Table. Distribution of Maternal Health Service Utilization Across Explanatory Variables.**

| **Variables** | **Weighted N ^a^** | **Weighted %^a^** | **ANC use % (95% CI) ^a^** | **p^b^** | **Adequate ANC % (95% CI) ^a^** | **p^b^** | **SBA % (95% CI) ^a^** | **p^b^** | **PNC % (95% CI) ^a^** | **p^b^** |
| --- | --- | --- | --- | --- | --- | --- | --- | --- | --- | --- |
| Residence |  |  |  |  |  |  |  |  |  |  |
| Urban | 2,149 | 47.7% | 92.1% (90.4–93.8) | <0.001 | 46.7% (43.4–50.1) | <0.001 | 94.9% (93.7–96.0) | <0.001 | 93.0% (91.4–94.6) | <0.001 |
| Rural | 2,358 | 52.3% | 85.3% (82.6–87.9) | <0.001 | 31.5% (28.4–34.5) | <0.001 | 79.3% (75.8–82.8) | <0.001 | 80.3% (77.0–83.5) | <0.001 |
| Phone type |  |  |  |  |  |  |  |  |  |  |
| No phone | 955 | 21.2% | 79.3% (75.4–83.2) | <0.001 | 28.0% (23.9–32.2) | <0.001 | 74.5% (69.9–79.0) | <0.001 | 76.1% (72.1–80.1) | <0.001 |
| Basic phone only | 1,989 | 44.1% | 87.7% (85.6–89.7) | <0.001 | 35.5% (32.5–38.5) | <0.001 | 86.0% (83.5–88.4) | <0.001 | 85.7% (83.3–88.1) | <0.001 |
| Smartphone | 1,563 | 34.7% | 95.2% (93.7–96.8) | <0.001 | 49.4% (46.0–52.9) | <0.001 | 95.1% (93.8–96.5) | <0.001 | 93.4% (91.8–94.9) | <0.001 |
| Internet use |  |  |  |  |  |  |  |  |  |  |
| Not regular | 3,180 | 70.6% | 85.8% (83.7–87.8) | <0.001 | 33.9% (31.2–36.6) | <0.001 | 82.6% (79.9–85.3) | <0.001 | 83.0% (80.6–85.5) | <0.001 |
| Regular | 1,327 | 29.4% | 95.1% (93.5–96.8) | <0.001 | 50.4% (46.7–54.0) | <0.001 | 96.6% (95.3–97.8) | <0.001 | 94.2% (92.5–95.8) | <0.001 |
| Newspaper |  |  |  |  |  |  |  |  |  |  |
| Not at all | 4,147 | 92.0% | 88.0% (86.3–89.7) | 0.003 | 37.5% (35.2–39.9) | <0.001 | 86.0% (83.9–88.1) | <0.001 | 85.7% (83.7–87.8) | 0.012 |
| Less than weekly | 261 | 5.8% | 92.7% (88.3–97.1) | 0.003 | 49.0% (41.3–56.8) | <0.001 | 94.9% (91.8–98.0) | <0.001 | 93.6% (90.1–97.2) | 0.012 |
| At least weekly | 98 | 2.2% | 98.6% (97.3–99.9) | 0.003 | 62.6% (50.2–75.1) | <0.001 | 95.6% (91.0–100.2) | <0.001 | 91.0% (83.0–98.9) | 0.012 |
| Radio |  |  |  |  |  |  |  |  |  |  |
| Not at all | 1,652 | 36.7% | 83.6% (80.7–86.5) | <0.001 | 31.6% (28.5–34.8) | <0.001 | 81.1% (77.1–85.2) | <0.001 | 80.7% (76.8–84.6) | <0.001 |
| Less than weekly | 1,073 | 23.8% | 91.1% (88.3–93.9) | <0.001 | 38.5% (34.0–43.0) | <0.001 | 90.0% (87.7–92.3) | <0.001 | 89.9% (87.8–92.1) | <0.001 |
| At least weekly | 1,781 | 39.5% | 91.5% (89.7–93.4) | <0.001 | 45.5% (42.2–48.7) | <0.001 | 89.9% (88.1–91.7) | <0.001 | 89.4% (87.5–91.3) | <0.001 |
| Television |  |  |  |  |  |  |  |  |  |  |
| Not at all | 1,351 | 30.0% | 81.2% (77.6–84.7) | <0.001 | 27.8% (24.4–31.2) | <0.001 | 75.0% (70.2–79.8) | <0.001 | 76.6% (72.2–81.0) | <0.001 |
| Less than weekly | 592 | 13.1% | 90.7% (87.5–93.9) | <0.001 | 38.1% (33.0–43.1) | <0.001 | 90.4% (87.7–93.1) | <0.001 | 88.4% (85.5–91.3) | <0.001 |
| At least weekly | 2,563 | 56.9% | 91.9% (90.3–93.5) | <0.001 | 44.7% (41.9–47.5) | <0.001 | 92.0% (90.5–93.5) | <0.001 | 91.0% (89.4–92.6) | <0.001 |
| Age group |  |  |  |  |  |  |  |  |  |  |
| 15-19 | 258 | 5.7% | 80.0% (73.9–86.2) | <0.001 | 28.5% (21.8–35.2) | 0.008 | 87.2% (82.7–91.8) | 0.096 | 87.2% (82.7–91.7) | 0.020 |
| 20-24 | 910 | 20.2% | 86.1% (83.1–89.1) | <0.001 | 34.7% (30.7–38.8) | 0.008 | 87.0% (84.0–89.9) | 0.096 | 85.4% (82.3–88.4) | 0.020 |
| 25-29 | 1,064 | 23.6% | 89.9% (87.4–92.3) | <0.001 | 38.8% (34.9–42.6) | 0.008 | 88.7% (86.4–91.0) | 0.096 | 88.8% (86.7–91.0) | 0.020 |
| 30-34 | 1,077 | 23.9% | 91.0% (88.8–93.2) | <0.001 | 43.4% (39.7–47.2) | 0.008 | 86.8% (83.7–89.8) | 0.096 | 87.0% (84.1–90.0) | 0.020 |
| 35-39 | 798 | 17.7% | 90.2% (87.6–92.8) | <0.001 | 40.3% (35.3–45.2) | 0.008 | 86.3% (83.3–89.4) | 0.096 | 85.8% (82.8–88.9) | 0.020 |
| 40-44 | 312 | 6.9% | 86.8% (81.6–92.0) | <0.001 | 38.0% (30.6–45.4) | 0.008 | 81.2% (75.0–87.3) | 0.096 | 79.6% (73.2–85.9) | 0.020 |
| 45-49 | 86 | 1.9% | 83.2% (74.2–92.3) | <0.001 | 41.2% (29.4–53.0) | 0.008 | 80.4% (71.3–89.5) | 0.096 | 82.3% (73.3–91.4) | 0.020 |
| Education |  |  |  |  |  |  |  |  |  |  |
| No education | 1,003 | 22.3% | 81.4% (77.3–85.4) | <0.001 | 27.8% (23.0–32.6) | <0.001 | 74.3% (68.6–80.0) | <0.001 | 75.6% (70.4–80.7) | <0.001 |
| Primary | 697 | 15.5% | 85.1% (82.0–88.3) | <0.001 | 31.0% (26.6–35.5) | <0.001 | 82.5% (78.9–86.1) | <0.001 | 82.9% (79.6–86.1) | <0.001 |
| Secondary | 2,387 | 53.0% | 90.9% (89.2–92.7) | <0.001 | 42.0% (39.2–44.8) | <0.001 | 91.0% (89.3–92.8) | <0.001 | 90.4% (88.6–92.1) | <0.001 |
| Higher | 419 | 9.3% | 97.6% (95.3–99.8) | <0.001 | 59.2% (53.1–65.3) | <0.001 | 98.8% (97.6–100.1) | <0.001 | 94.7% (91.8–97.6) | <0.001 |
| Religion |  |  |  |  |  |  |  |  |  |  |
| Christian | 3,130 | 69.5% | 89.1% (87.4–90.8) | <0.001 | 41.5% (38.9–44.1) | <0.001 | 88.6% (86.8–90.3) | <0.001 | 88.1% (86.3–89.9) | <0.001 |
| Muslim | 1,126 | 25.0% | 91.3% (89.3–93.4) | <0.001 | 36.7% (32.4–41.0) | <0.001 | 89.3% (87.1–91.5) | <0.001 | 87.5% (85.2–89.8) | <0.001 |
| Traditional | 145 | 3.2% | 62.4% (50.5–74.3) | <0.001 | 6.1% (2.1–10.2) | <0.001 | 39.8% (22.7–57.0) | <0.001 | 44.1% (27.8–60.3) | <0.001 |
| No religion/Other | 105 | 2.3% | 76.7% (68.0–85.4) | <0.001 | 23.7% (13.2–34.1) | <0.001 | 69.0% (58.1–79.8) | <0.001 | 80.1% (69.8–90.4) | <0.001 |
| Marital status |  |  |  |  |  |  |  |  |  |  |
| Never married | 525 | 11.6% | 85.6% (81.4–89.9) | <0.001 | 36.5% (30.6–42.4) | 0.171 | 90.9% (87.6–94.2) | 0.015 | 90.2% (86.5–93.8) | 0.090 |
| Married | 2,780 | 61.7% | 90.8% (89.0–92.6) | <0.001 | 40.2% (37.4–43.0) | 0.171 | 87.4% (84.8–89.9) | 0.015 | 86.7% (84.3–89.0) | 0.090 |
| Cohabiting | 975 | 21.6% | 85.4% (82.3–88.5) | <0.001 | 37.6% (33.4–41.9) | 0.171 | 83.5% (80.3–86.6) | 0.015 | 84.0% (80.5–87.5) | 0.090 |
| Formerly married | 227 | 5.0% | 81.0% (74.4–87.6) | <0.001 | 31.2% (23.1–39.2) | 0.171 | 82.8% (76.6–89.1) | 0.015 | 83.2% (76.5–89.8) | 0.090 |
| Working status |  |  |  |  |  |  |  |  |  |  |
| Not working | 991 | 22.0% | 85.6% (82.5–88.7) | 0.016 | 33.9% (30.1–37.8) | 0.006 | 88.8% (86.3–91.3) | 0.086 | 87.2% (84.5–90.0) | 0.472 |
| Working | 3,516 | 78.0% | 89.4% (87.6–91.1) | 0.016 | 40.1% (37.5–42.7) | 0.006 | 86.1% (83.8–88.4) | 0.086 | 86.1% (83.9–88.2) | 0.472 |
| Parity |  |  |  |  |  |  |  |  |  |  |
| None | 31 | 0.7% | 82.3% (66.3–98.3) | 0.005 | 21.9% (-1.6–45.5) | 0.007 | 72.5% (54.6–90.4) | <0.001 | 68.5% (43.1–93.8) | <0.001 |
| One | 1,210 | 26.9% | 90.4% (88.2–92.5) | 0.005 | 40.9% (37.2–44.5) | 0.007 | 92.5% (90.7–94.4) | <0.001 | 90.8% (88.8–92.8) | <0.001 |
| Two | 1,015 | 22.5% | 90.5% (88.1–92.8) | 0.005 | 43.2% (39.3–47.0) | 0.007 | 89.7% (87.5–92.0) | <0.001 | 89.1% (86.9–91.3) | <0.001 |
| Three | 750 | 16.6% | 89.0% (86.5–91.6) | 0.005 | 38.8% (34.2–43.4) | 0.007 | 88.6% (86.0–91.2) | <0.001 | 87.4% (84.7–90.2) | <0.001 |
| Four or more | 1,501 | 33.3% | 85.6% (82.9–88.4) | 0.005 | 34.4% (30.4–38.4) | 0.007 | 79.3% (75.4–83.2) | <0.001 | 80.7% (76.9–84.4) | <0.001 |
| Health insurance |  |  |  |  |  |  |  |  |  |  |
| Not insured | 227 | 5.0% | 67.2% (57.9–76.5) | <0.001 | 23.1% (16.0–30.2) | <0.001 | 64.7% (56.1–73.4) | <0.001 | 65.3% (57.2–73.4) | <0.001 |
| Insured | 4,280 | 95.0% | 89.7% (88.2–91.1) | <0.001 | 39.6% (37.3–41.9) | <0.001 | 87.9% (86.1–89.7) | <0.001 | 87.4% (85.7–89.2) | <0.001 |
| Wealth index |  |  |  |  |  |  |  |  |  |  |
| Poorest | 1,071 | 23.8% | 78.7% (74.2–83.3) | <0.001 | 23.4% (19.7–27.1) | <0.001 | 72.4% (66.8–78.0) | <0.001 | 73.2% (68.1–78.3) | <0.001 |
| Poorer | 948 | 21.0% | 88.9% (86.6–91.1) | <0.001 | 34.5% (29.8–39.1) | <0.001 | 84.9% (81.2–88.6) | <0.001 | 84.8% (81.4–88.3) | <0.001 |
| Middle | 882 | 19.6% | 86.5% (82.5–90.4) | <0.001 | 34.8% (30.5–39.1) | <0.001 | 88.2% (84.9–91.4) | <0.001 | 87.7% (84.3–91.0) | <0.001 |
| Richer | 822 | 18.2% | 94.6% (92.4–96.7) | <0.001 | 47.6% (43.1–52.2) | <0.001 | 95.6% (93.9–97.4) | <0.001 | 94.9% (92.8–96.9) | <0.001 |
| Richest | 783 | 17.4% | 97.5% (95.8–99.2) | <0.001 | 60.1% (54.4–65.7) | <0.001 | 97.4% (95.9–99.0) | <0.001 | 95.6% (93.5–97.6) | <0.001 |
| Electricity |  |  |  |  |  |  |  |  |  |  |
| No electricity | 894 | 19.8% | 77.5% (72.2–82.8) | <0.001 | 24.6% (20.5–28.7) | <0.001 | 72.0% (65.8–78.3) | <0.001 | 73.6% (67.9–79.3) | <0.001 |
| Has electricity | 3,613 | 80.2% | 91.3% (89.9–92.6) | <0.001 | 42.2% (39.7–44.8) | <0.001 | 90.3% (88.5–92.2) | <0.001 | 89.5% (87.6–91.3) | <0.001 |
| Any barrier |  |  |  |  |  |  |  |  |  |  |
| No barrier | 2,001 | 44.4% | 92.5% (90.8–94.3) | <0.001 | 46.1% (43.0–49.3) | <0.001 | 92.1% (90.7–93.6) | <0.001 | 90.9% (89.3–92.6) | <0.001 |
| Any barrier | 2,505 | 55.6% | 85.3% (83.1–87.6) | <0.001 | 32.9% (29.9–35.8) | <0.001 | 82.4% (79.4–85.3) | <0.001 | 82.6% (79.8–85.5) | <0.001 |

^a^Weighted N and % use DHS sampling weights (V005/1,000,000). Prevalences are survey-weighted percentages with 95% confidence intervals in parentheses.

^b^p-values from Rao-Scott adjusted F test accounting for complex survey design.
