## Supplementary table for "Digital access, media exposure, and maternal health service utilization among women in Ghana"

**S5 Table. Unadjusted Odds Ratios - Media Exposure and Maternal Health Service Utilization (Model I, Total Sample, N = 4,982)**

| **Variables** | **ANC use (4+)** | **Adequate ANC (8+)** | **SBA** | **PNC 6 weeks** |
| --- | --- | --- | --- | --- |
| **Frequency of reading newspaper** | | | | |
| Not at all | 1 [Ref.] | 1 [Ref.] | 1 [Ref.] | 1 [Ref.] |
| Less than once a week | 1.28 [0.74,2.20] | 1.36 [1.01,1.81]* | 1.74 [0.95,3.17] | 1.49 [0.87,2.58] |
| At least once a week | 1.95 [0.70,5.49] | 1.57 [1.00,2.46]* | 1.63 [0.63,4.25] | 1.15 [0.52,2.55] |
| **Frequency of listening to radio** | | | | |
| Not at all | 1 [Ref.] | 1 [Ref.] | 1 [Ref.] | 1 [Ref.] |
| Less than once a week | 1.37 [1.04,1.81]* | 1.01 [0.85,1.21] | 1.19 [0.91,1.56] | 1.18 [0.91,1.53] |
| At least once a week | 1.22 [0.97,1.52] | 1.30 [1.12,1.52]*** | 1.09 [0.88,1.36] | 1.06 [0.85,1.31] |
| **Frequency of watching television** | | | | |
| Not at all | 1 [Ref.] | 1 [Ref.] | 1 [Ref.] | 1 [Ref.] |
| Less than once a week | 1.87 [1.35,2.59]*** | 1.48 [1.20,1.83]*** | 1.89 [1.38,2.59]*** | 1.31 [0.98,1.76] |
| At least once a week | 1.95 [1.56,2.44]*** | 1.71 [1.46,2.01]*** | 2.07 [1.66,2.60]*** | 1.71 [1.38,2.14]*** |
| **Random Effects** | | | | |
| PSU variance | 0.8746 | 0.5956 | 1.3860 | 1.4539 |
| ICC | 21.0% | 15.3% | 29.6% | 30.6% |
| MOR | 2.440 | 2.088 | 3.074 | 3.159 |
| **Model Fitness** | | | | |
| Log-likelihood | -1710.37 | -3054.21 | -1794.35 | -1885.21 |
| AIC | 3436.73 | 6124.42 | 3604.71 | 3786.42 |
| Wald chi-square | — | — | — | — |
| N | 4,982 | 4,982 | 4,982 | 4,982 |
| Number of clusters | 617 | 617 | 617 | 617 |

aOR = adjusted odds ratio; 95% CI = 95% confidence intervals. * p<0.05  ** p<0.01  *** p<0.001. Ref. = Reference category. PSU = Primary Sampling Unit. ICC = Intra-Class Correlation. MOR = Median Odds Ratio. AIC = Akaike Information Criterion. Model I: exposure variables only, no covariates.
