## Supplementary table for "Digital access, media exposure, and maternal health service utilization among women in Ghana"

**S6 Table. Unadjusted Odds Ratio for Media Exposure - Urban and Rural Strata**

| **Variables** | **ANC use (4+)** | **Adequate ANC (8+)** | **SBA** | **PNC 6 weeks** |
| --- | --- | --- | --- | --- |
| **Urban Stratum (N = 2,046)** | | | | |
| **Frequency of reading newspaper** | | | | |
| Not at all | 1 [Ref.] | 1 [Ref.] | 1 [Ref.] | 1 [Ref.] |
| Less than once a week | 1.66 [0.71,3.92] | 1.22 [0.84,1.77] | 3.61 [0.87,14.97] | 2.64 [0.93,7.46] |
| At least once a week | 2.17 [0.51,9.20] | 1.62 [0.93,2.83] | 1.62 [0.38,6.91] | 0.84 [0.31,2.24] |
| **Frequency of listening to radio** | | | | |
| Not at all | 1 [Ref.] | 1 [Ref.] | 1 [Ref.] | 1 [Ref.] |
| Less than once a week | 1.14 [0.73,1.78] | 0.99 [0.77,1.27] | 1.34 [0.80,2.25] | 1.08 [0.68,1.72] |
| At least once a week | 1.16 [0.78,1.73] | 1.31 [1.05,1.65]* | 1.32 [0.84,2.09] | 1.07 [0.71,1.63] |
| **Frequency of watching television** | | | | |
| Not at all | 1 [Ref.] | 1 [Ref.] | 1 [Ref.] | 1 [Ref.] |
| Less than once a week | 1.46 [0.84,2.55] | 1.55 [1.10,2.19]* | 1.67 [0.84,3.30] | 1.23 [0.67,2.22] |
| At least once a week | 1.90 [1.27,2.86]** | 1.62 [1.23,2.12]*** | 1.46 [0.91,2.34] | 1.31 [0.85,2.03] |
| **Random Effects** | | | | |
| PSU variance | 0.5950 | 0.3583 | 0.5078 | 0.9888 |
| ICC | 15.3% | 9.8% | 13.4% | 23.1% |
| MOR | 2.087 | 1.770 | 1.973 | 2.582 |
| **Model Fitness** | | | | |
| Log-likelihood | -556.41 | -1362.66 | -436.99 | -540.77 |
| AIC | 1128.81 | 2741.32 | 889.99 | 1097.53 |
| Wald chi-square | — | — | — | — |
| N | 2,046 | 2,046 | 2,046 | 2,046 |
| Number of clusters | 303 | 303 | 303 | 303 |
| **Rural Stratum (N = 2,936)** | | | | |
| **Frequency of reading newspaper** | | | | |
| Not at all | 1 [Ref.] | 1 [Ref.] | 1 [Ref.] | 1 [Ref.] |
| Less than once a week | 1.01 [0.49,2.06] | 1.57 [1.00,2.46]* | 1.29 [0.65,2.57] | 1.06 [0.55,2.07] |
| At least once a week | 1.62 [0.36,7.17] | 1.32 [0.62,2.82] | 1.37 [0.38,4.92] | 1.59 [0.42,5.95] |
| **Frequency of listening to radio** | | | | |
| Not at all | 1 [Ref.] | 1 [Ref.] | 1 [Ref.] | 1 [Ref.] |
| Less than once a week | 1.50 [1.05,2.14]* | 1.00 [0.78,1.28] | 1.12 [0.82,1.54] | 1.21 [0.88,1.66] |
| At least once a week | 1.23 [0.94,1.60] | 1.29 [1.05,1.59]* | 1.06 [0.83,1.37] | 1.06 [0.82,1.36] |
| **Frequency of watching television** | | | | |
| Not at all | 1 [Ref.] | 1 [Ref.] | 1 [Ref.] | 1 [Ref.] |
| Less than once a week | 2.07 [1.36,3.15]*** | 1.30 [0.99,1.72] | 1.73 [1.21,2.48]** | 1.21 [0.86,1.71] |
| At least once a week | 1.71 [1.29,2.26]*** | 1.50 [1.21,1.85]*** | 1.87 [1.43,2.44]*** | 1.60 [1.23,2.10]*** |
| **Random Effects** | | | | |
| PSU variance | 0.9588 | 0.7463 | 1.3893 | 1.4572 |
| ICC | 22.6% | 18.5% | 29.7% | 30.7% |
| MOR | 2.545 | 2.280 | 3.078 | 3.163 |
| **Model Fitness** | | | | |
| Log-likelihood | -1146.42 | -1672.63 | -1320.67 | -1322.78 |
| AIC | 2308.84 | 3361.26 | 2657.34 | 2661.56 |
| Wald chi-square | — | — | — | — |
| N | 2,936 | 2,936 | 2,936 | 2,936 |
| Number of clusters | 314 | 314 | 314 | 314 |

aOR = adjusted odds ratio; 95% CI = 95% confidence intervals. * p<0.05  ** p<0.01  *** p<0.001. Ref. = Reference category. PSU = Primary Sampling Unit. ICC = Intra-Class Correlation. MOR = Median Odds Ratio. AIC = Akaike Information Criterion.
