## Supplementary table for "Digital access, media exposure, and maternal health service utilization among women in Ghana"

**S7 Table. Unadjusted Odds Ratios - Digital Access and Maternal Health Service Utilization (Model I, Total Sample, N = 4,982)**

| **Variables** | **ANC use (4+)** | **Adequate ANC (8+)** | **SBA** | **PNC 6 weeks** |
| --- | --- | --- | --- | --- |
| **Phone type** | | | | |
| No phone | 1 [Ref.] | 1 [Ref.] | 1 [Ref.] | 1 [Ref.] |
| Basic phone only | 1.74 [1.42,2.13]*** | 1.26 [1.07,1.49]** | 1.75 [1.43,2.13]*** | 1.68 [1.37,2.05]*** |
| Smartphone | 3.32 [2.22,4.97]*** | 1.84 [1.45,2.33]*** | 2.68 [1.84,3.92]*** | 2.06 [1.45,2.91]*** |
| **Frequency of internet use** | | | | |
| Not regular | 1 [Ref.] | 1 [Ref.] | 1 [Ref.] | 1 [Ref.] |
| Regular user | 1.38 [0.90,2.10] | 1.28 [1.03,1.59]* | 2.07 [1.35,3.17]*** | 1.55 [1.07,2.26]* |
| **Random Effects** | | | | |
| PSU variance | 0.8816 | 0.6114 | 1.2963 | 1.4232 |
| ICC | 21.1% | 15.7% | 28.3% | 30.2% |
| MOR | 2.449 | 2.108 | 2.963 | 3.120 |
| **Model Fitness** | | | | |
| Log-likelihood | -1690.97 | -3053.83 | -1769.54 | -1869.78 |
| AIC | 3391.94 | 6117.65 | 3549.08 | 3749.56 |
| Wald chi-square | — | — | — | — |
| N | 4,982 | 4,982 | 4,982 | 4,982 |
| Number of clusters | 617 | 617 | 617 | 617 |

aOR = adjusted odds ratio; 95% CI = 95% confidence intervals. * p<0.05  ** p<0.01  *** p<0.001. Ref. = Reference category. PSU = Primary Sampling Unit. ICC = Intra-Class Correlation. MOR = Median Odds Ratio. AIC = Akaike Information Criterion. Model I: exposure variables only.
