## Supplementary table for "Digital access, media exposure, and maternal health service utilization among women in Ghana"

**S8 Table. Unadjusted Odds Ratios for Digital Access - Urban and Rural Strata**

| **Variables** | **ANC use (4+)** | **Adequate ANC (8+)** | **SBA** | **PNC 6 weeks** |
| --- | --- | --- | --- | --- |
| **Urban Stratum (N = 2,046)** | | | | |
| **Phone type** | | | | |
| No phone | 1 [Ref.] | 1 [Ref.] | 1 [Ref.] | 1 [Ref.] |
| Basic phone only | 1.53 [1.01,2.33]* | 1.10 [0.82,1.49] | 2.02 [1.28,3.18]** | 1.45 [0.92,2.30] |
| Smartphone | 4.08 [2.14,7.80]*** | 1.74 [1.21,2.49]** | 3.51 [1.72,7.14]*** | 2.05 [1.10,3.82]* |
| **Frequency of internet use** | | | | |
| Not regular | 1 [Ref.] | 1 [Ref.] | 1 [Ref.] | 1 [Ref.] |
| Regular user | 1.19 [0.67,2.12] | 1.03 [0.78,1.36] | 2.03 [1.00,4.10]* | 1.30 [0.75,2.25] |
| **Random Effects** | | | | |
| PSU variance | 0.5715 | 0.3546 | 0.4104 | 0.9455 |
| ICC | 14.8% | 9.7% | 11.1% | 22.3% |
| MOR | 2.057 | 1.765 | 1.842 | 2.528 |
| **Model Fitness** | | | | |
| Log-likelihood | -540.82 | -1364.06 | -421.06 | -536.64 |
| AIC | 1091.65 | 2738.11 | 852.12 | 1083.28 |
| Wald chi-square | — | — | — | — |
| N | 2,046 | 2,046 | 2,046 | 2,046 |
| Number of clusters | 303 | 303 | 303 | 303 |
| **Rural Stratum (N = 2,936)** | | | | |
| **Phone type** | | | | |
| No phone | 1 [Ref.] | 1 [Ref.] | 1 [Ref.] | 1 [Ref.] |
| Basic phone only | 1.76 [1.39,2.23]*** | 1.25 [1.02,1.53]* | 1.60 [1.28,1.99]*** | 1.65 [1.32,2.06]*** |
| Smartphone | 2.48 [1.47,4.16]*** | 1.55 [1.11,2.17]** | 2.14 [1.36,3.35]*** | 1.73 [1.12,2.67]* |
| **Frequency of internet use** | | | | |
| Not regular | 1 [Ref.] | 1 [Ref.] | 1 [Ref.] | 1 [Ref.] |
| Regular user | 1.31 [0.70,2.45] | 1.58 [1.11,2.25]* | 1.57 [0.91,2.72] | 1.48 [0.87,2.51] |
| **Random Effects** | | | | |
| PSU variance | 0.9835 | 0.7743 | 1.3610 | 1.4707 |
| ICC | 23.0% | 19.1% | 29.3% | 30.9% |
| MOR | 2.575 | 2.315 | 3.043 | 3.180 |
| **Model Fitness** | | | | |
| Log-likelihood | -1144.61 | -1671.04 | -1316.23 | -1315.85 |
| AIC | 2299.21 | 3352.09 | 2642.47 | 2641.70 |
| Wald chi-square | — | — | — | — |
| N | 2,936 | 2,936 | 2,936 | 2,936 |
| Number of clusters | 314 | 314 | 314 | 314 |

aOR = adjusted odds ratio; 95% CI = 95% confidence intervals. * p<0.05  ** p<0.01  *** p<0.001. Ref. = Reference category. PSU = Primary Sampling Unit. ICC = Intra-Class Correlation. MOR = Median Odds Ratio. AIC = Akaike Information Criterion.
