## Supplementary table for "Digital access, media exposure, and maternal health service utilization among women in Ghana"

**S9 Table. Fully Adjusted Odds Ratios for All Variables - Media Model II (Total Sample, N = 4,982)**

| **Variables** | **ANC use (4+)** | **Adequate ANC (8+)** | **SBA** | **PNC 6 weeks** |
| --- | --- | --- | --- | --- |
| **Exposure Variables** | | | | |
| **Frequency of reading newspaper** | | | | |
| Not at all | 1 [Ref.] | 1 [Ref.] | 1 [Ref.] | 1 [Ref.] |
| Less than once a week | 0.97 [0.54,1.72] | 1.09 [0.81,1.47] | 1.19 [0.64,2.23] | 1.15 [0.66,2.02] |
| At least once a week | 1.22 [0.41,3.61] | 1.14 [0.71,1.83] | 0.85 [0.30,2.38] | 0.81 [0.35,1.89] |
| **Frequency of listening to radio** | | | | |
| Not at all | 1 [Ref.] | 1 [Ref.] | 1 [Ref.] | 1 [Ref.] |
| Less than once a week | 1.17 [0.88,1.56] | 0.90 [0.75,1.09] | 1.05 [0.79,1.38] | 1.06 [0.81,1.39] |
| At least once a week | 1.13 [0.89,1.42] | 1.23 [1.05,1.44]* | 1.10 [0.87,1.38] | 1.03 [0.83,1.29] |
| **Frequency of watching television** | | | | |
| Not at all | 1 [Ref.] | 1 [Ref.] | 1 [Ref.] | 1 [Ref.] |
| Less than once a week | 1.36 [0.96,1.92] | 1.05 [0.84,1.32] | 1.26 [0.90,1.75] | 0.96 [0.70,1.30] |
| At least once a week | 1.15 [0.89,1.50] | 1.02 [0.84,1.24] | 1.15 [0.89,1.49] | 1.11 [0.86,1.43] |
| **Covariates** | | | | |
| **Age group (years)** | | | | |
| 15–19 | 1 [Ref.] | 1 [Ref.] | 1 [Ref.] | 1 [Ref.] |
| 20–24 | 1.63 [1.10,2.41]* | 1.46 [1.05,2.04]* | 1.02 [0.65,1.61] | 1.08 [0.71,1.65] |
| 25–29 | 2.52 [1.60,3.98]*** | 1.70 [1.20,2.43]** | 1.58 [0.95,2.63] | 1.49 [0.93,2.39] |
| 30–34 | 3.06 [1.83,5.10]*** | 2.12 [1.45,3.11]*** | 1.63 [0.94,2.83] | 1.53 [0.91,2.57] |
| 35–39 | 2.69 [1.55,4.65]*** | 1.84 [1.23,2.77]** | 1.57 [0.88,2.82] | 1.53 [0.88,2.67] |
| 40–44 | 3.37 [1.82,6.24]*** | 1.97 [1.25,3.11]** | 1.79 [0.94,3.39] | 1.36 [0.75,2.49] |
| 45–49 | 2.35 [1.09,5.07]* | 1.85 [0.98,3.48] | 1.74 [0.78,3.90] | 1.84 [0.84,4.03] |
| **Education level** | | | | |
| No education | 1 [Ref.] | 1 [Ref.] | 1 [Ref.] | 1 [Ref.] |
| Primary | 1.10 [0.83,1.47] | 0.96 [0.76,1.20] | 1.03 [0.78,1.35] | 0.86 [0.66,1.12] |
| Secondary | 1.23 [0.93,1.62] | 1.05 [0.86,1.28] | 1.33 [1.01,1.74]* | 1.10 [0.85,1.43] |
| Higher | 2.45 [0.91,6.60] | 1.20 [0.86,1.67] | 2.01 [0.79,5.13] | 1.03 [0.53,2.01] |
| **Religion** | | | | |
| Christian | 1 [Ref.] | 1 [Ref.] | 1 [Ref.] | 1 [Ref.] |
| Muslim | 1.10 [0.83,1.46] | 1.05 [0.87,1.26] | 0.90 [0.68,1.19] | 1.11 [0.84,1.46] |
| Traditional | 0.57 [0.35,0.94]* | 0.35 [0.17,0.69]** | 0.32 [0.19,0.52]*** | 0.41 [0.25,0.68]*** |
| No religion/Other | 0.69 [0.41,1.14] | 0.51 [0.29,0.87]* | 0.54 [0.33,0.89]* | 0.80 [0.49,1.33] |
| **Marital status** | | | | |
| Never married | 1 [Ref.] | 1 [Ref.] | 1 [Ref.] | 1 [Ref.] |
| Married | 1.70 [1.18,2.46]** | 1.16 [0.89,1.52] | 1.09 [0.72,1.65] | 0.95 [0.64,1.42] |
| Cohabiting | 1.13 [0.78,1.63] | 1.13 [0.86,1.48] | 0.77 [0.51,1.17] | 0.74 [0.49,1.10] |
| Formerly married | 0.91 [0.54,1.51] | 0.89 [0.60,1.31] | 0.72 [0.41,1.25] | 0.71 [0.41,1.22] |
| **Working status** | | | | |
| Not working | 1 [Ref.] | 1 [Ref.] | 1 [Ref.] | 1 [Ref.] |
| Working | 1.23 [0.98,1.54] | 1.09 [0.92,1.28] | 0.79 [0.62,1.00]* | 0.80 [0.63,1.01] |
| **Parity** | | | | |
| One | 1 [Ref.] | 1 [Ref.] | 1 [Ref.] | 1 [Ref.] |
| Two | 0.77 [0.55,1.07] | 0.87 [0.71,1.06] | 0.64 [0.46,0.89]** | 0.75 [0.55,1.03] |
| Three | 0.47 [0.32,0.69]*** | 0.70 [0.55,0.88]** | 0.47 [0.32,0.69]*** | 0.60 [0.42,0.86]** |
| Four or more | 0.45 [0.29,0.68]*** | 0.66 [0.51,0.85]** | 0.39 [0.26,0.58]*** | 0.55 [0.37,0.80]** |
| **Health insurance** | | | | |
| Not insured | 1 [Ref.] | 1 [Ref.] | 1 [Ref.] | 1 [Ref.] |
| Insured | 2.38 [1.71,3.30]*** | 1.42 [1.00,2.00]* | 2.33 [1.67,3.25]*** | 2.35 [1.70,3.25]*** |
| **Wealth index** | | | | |
| Poorest | 1 [Ref.] | 1 [Ref.] | 1 [Ref.] | 1 [Ref.] |
| Poorer | 1.44 [1.04,1.99]* | 1.29 [1.00,1.66]* | 1.18 [0.86,1.61] | 1.05 [0.78,1.41] |
| Middle | 1.58 [1.03,2.44]* | 1.34 [0.98,1.82] | 1.52 [0.99,2.33] | 1.27 [0.84,1.92] |
| Richer | 2.92 [1.68,5.06]*** | 1.53 [1.09,2.15]* | 2.98 [1.70,5.24]*** | 2.37 [1.40,4.00]** |
| Richest | 5.23 [2.35,11.63]*** | 2.36 [1.60,3.49]*** | 3.28 [1.52,7.08]** | 2.43 [1.25,4.73]** |
| **Place of residence** | | | | |
| Rural | 1 [Ref.] | 1 [Ref.] | 1 [Ref.] | 1 [Ref.] |
| Urban | 0.88 [0.66,1.17] | 1.14 [0.93,1.39] | 1.93 [1.42,2.63]*** | 1.47 [1.08,2.00]* |
| **Electricity** | | | | |
| No electricity | 1 [Ref.] | 1 [Ref.] | 1 [Ref.] | 1 [Ref.] |
| Has electricity | 1.37 [0.99,1.89] | 1.20 [0.92,1.57] | 1.46 [1.06,2.01]* | 1.42 [1.03,1.96]* |
| **Region** | | | | |
| Western | 1 [Ref.] | 1 [Ref.] | 1 [Ref.] | 1 [Ref.] |
| Central | 0.66 [0.31,1.44] | 0.68 [0.42,1.12] | 0.86 [0.43,1.70] | 1.00 [0.48,2.07] |
| Greater Accra | 0.35 [0.16,0.78]* | 0.61 [0.37,1.00] | 0.99 [0.45,2.16] | 1.22 [0.54,2.76] |
| Volta | 1.23 [0.51,3.01] | 0.58 [0.34,0.98]* | 2.75 [1.21,6.25]* | 1.40 [0.64,3.07] |
| Eastern | 0.68 [0.30,1.54] | 0.77 [0.46,1.28] | 1.79 [0.81,3.93] | 1.86 [0.81,4.28] |
| Ashanti | 0.75 [0.35,1.62] | 0.64 [0.39,1.03] | 2.17 [1.03,4.53]* | 1.29 [0.62,2.69] |
| Western North | 0.63 [0.29,1.36] | 0.41 [0.24,0.70]*** | 2.19 [1.04,4.62]* | 1.20 [0.57,2.53] |
| Ahafo | 1.14 [0.51,2.54] | 0.92 [0.56,1.53] | 4.35 [1.95,9.71]*** | 4.39 [1.83,10.56]*** |
| Bono | 0.83 [0.36,1.91] | 0.55 [0.33,0.93]* | 2.14 [0.97,4.72] | 2.54 [1.07,6.01]* |
| Bono East | 0.83 [0.39,1.76] | 0.69 [0.42,1.13] | 1.58 [0.79,3.13] | 1.24 [0.60,2.55] |
| Oti | 0.51 [0.25,1.05] | 0.47 [0.28,0.78]** | 0.91 [0.47,1.74] | 0.99 [0.49,1.99] |
| Northern | 0.66 [0.31,1.40] | 0.41 [0.25,0.68]*** | 1.36 [0.69,2.68] | 0.76 [0.38,1.54] |
| Savannah | 0.70 [0.33,1.46] | 0.41 [0.25,0.69]*** | 1.29 [0.66,2.51] | 0.83 [0.41,1.67] |
| North East | 0.60 [0.28,1.26] | 0.44 [0.26,0.73]** | 1.87 [0.94,3.72] | 1.00 [0.49,2.05] |
| Upper East | 2.43 [1.00,5.90] | 0.83 [0.51,1.36] | 13.64 [5.10,36.44]*** | 4.90 [2.08,11.52]*** |
| Upper West | 1.93 [0.81,4.57] | 0.55 [0.33,0.91]* | 3.04 [1.45,6.38]** | 1.86 [0.87,4.00] |
| **Any barrier to healthcare** | | | | |
| No barrier | 1 [Ref.] | 1 [Ref.] | 1 [Ref.] | 1 [Ref.] |
| Any barrier | 0.81 [0.65,1.01] | 0.73 [0.63,0.84]*** | 0.86 [0.69,1.07] | 0.91 [0.74,1.13] |
| **Random Effects** | | | | |
| PSU variance | 0.4776 | 0.4349 | 0.5646 | 0.8200 |
| ICC | 12.7% | 11.7% | 14.6% | 20.0% |
| MOR | 1.933 | 1.876 | 2.048 | 2.372 |
| **Model Fitness** | | | | |
| Log-likelihood | -1570.11 | -2925.66 | -1609.18 | -1760.80 |
| AIC | 3240.23 | 5951.33 | 3318.36 | 3621.60 |
| Wald chi-square | 346.94*** | 343.85*** | 438.90*** | 286.49*** |
| N | 4,982 | 4,982 | 4,982 | 4,982 |
| Number of clusters | 617 | 617 | 617 | 617 |

aOR = adjusted odds ratio; 95% CI = 95% confidence intervals. * p<0.05  ** p<0.01  *** p<0.001. Ref. = Reference category. PSU = Primary Sampling Unit. ICC = Intra-Class Correlation. MOR = Median Odds Ratio. AIC = Akaike Information Criterion. Region entered as a 16-category factor (Western Region as reference) per convention in Ghana DHS multilevel analyses. Parity "None" (n=32) merged into "One" before modelling due to extreme collinearity (VIF=38.7) arising from the near-empty reference category.
