## Supplementary table for "Digital access, media exposure, and maternal health service utilization among women in Ghana"

**S10 Table. Fully Adjusted Odds Ratios for All Variables - Digital Model II (Total Sample, N = 4,982)**

| **Variables** | **ANC use (4+)** | **Adequate ANC (8+)** | **SBA** | **PNC 6 weeks** |
| --- | --- | --- | --- | --- |
| **Exposure Variables** | | | | |
| **Phone type** | | | | |
| No phone | 1 [Ref.] | 1 [Ref.] | 1 [Ref.] | 1 [Ref.] |
| Basic phone only | 1.37 [1.11,1.70]** | 1.02 [0.86,1.22] | 1.41 [1.15,1.73]*** | 1.38 [1.12,1.69]** |
| Smartphone | 1.97 [1.30,2.97]** | 1.26 [0.98,1.62] | 1.50 [1.01,2.22]* | 1.27 [0.89,1.83] |
| **Frequency of internet use** | | | | |
| Not regular | 1 [Ref.] | 1 [Ref.] | 1 [Ref.] | 1 [Ref.] |
| Regular user | 0.89 [0.57,1.38] | 0.88 [0.70,1.12] | 1.21 [0.77,1.91] | 1.05 [0.70,1.57] |
| **Covariates** | | | | |
| **Age group (years)** | | | | |
| 15–19 | 1 [Ref.] | 1 [Ref.] | 1 [Ref.] | 1 [Ref.] |
| 20–24 | 1.54 [1.04,2.28]* | 1.44 [1.03,2.01]* | 0.96 [0.61,1.52] | 1.04 [0.68,1.59] |
| 25–29 | 2.36 [1.50,3.74]*** | 1.67 [1.17,2.38]** | 1.47 [0.89,2.45] | 1.42 [0.88,2.28] |
| 30–34 | 2.78 [1.66,4.66]*** | 2.09 [1.42,3.06]*** | 1.48 [0.85,2.58] | 1.42 [0.84,2.40] |
| 35–39 | 2.46 [1.42,4.26]** | 1.82 [1.21,2.73]** | 1.43 [0.80,2.56] | 1.43 [0.82,2.50] |
| 40–44 | 3.12 [1.69,5.79]*** | 1.98 [1.25,3.12]** | 1.63 [0.86,3.09] | 1.27 [0.69,2.32] |
| 45–49 | 2.18 [1.01,4.73]* | 1.80 [0.96,3.38] | 1.59 [0.71,3.57] | 1.70 [0.77,3.73] |
| **Education level** | | | | |
| No education | 1 [Ref.] | 1 [Ref.] | 1 [Ref.] | 1 [Ref.] |
| Primary | 1.09 [0.82,1.46] | 0.96 [0.77,1.20] | 1.02 [0.77,1.33] | 0.86 [0.65,1.12] |
| Secondary | 1.18 [0.90,1.55] | 1.06 [0.87,1.30] | 1.28 [0.98,1.67] | 1.08 [0.83,1.40] |
| Higher | 2.19 [0.80,5.96] | 1.24 [0.88,1.73] | 1.71 [0.66,4.40] | 0.99 [0.50,1.95] |
| **Religion** | | | | |
| Christian | 1 [Ref.] | 1 [Ref.] | 1 [Ref.] | 1 [Ref.] |
| Muslim | 1.08 [0.81,1.43] | 1.02 [0.85,1.23] | 0.87 [0.66,1.15] | 1.09 [0.83,1.44] |
| Traditional | 0.57 [0.35,0.93]* | 0.34 [0.17,0.67]** | 0.31 [0.19,0.51]*** | 0.42 [0.25,0.69]*** |
| No religion/Other | 0.68 [0.41,1.13] | 0.50 [0.29,0.86]* | 0.54 [0.33,0.88]* | 0.81 [0.49,1.35] |
| **Marital status** | | | | |
| Never married | 1 [Ref.] | 1 [Ref.] | 1 [Ref.] | 1 [Ref.] |
| Married | 1.70 [1.18,2.47]** | 1.18 [0.91,1.53] | 1.12 [0.74,1.69] | 0.97 [0.65,1.44] |
| Cohabiting | 1.14 [0.79,1.65] | 1.14 [0.87,1.50] | 0.80 [0.53,1.20] | 0.75 [0.50,1.12] |
| Formerly married | 0.89 [0.53,1.48] | 0.88 [0.59,1.30] | 0.72 [0.41,1.24] | 0.71 [0.41,1.23] |
| **Working status** | | | | |
| Not working | 1 [Ref.] | 1 [Ref.] | 1 [Ref.] | 1 [Ref.] |
| Working | 1.24 [0.99,1.56] | 1.11 [0.94,1.30] | 0.79 [0.62,1.00] | 0.80 [0.63,1.00] |
| **Parity** | | | | |
| One | 1 [Ref.] | 1 [Ref.] | 1 [Ref.] | 1 [Ref.] |
| Two | 0.79 [0.57,1.11] | 0.88 [0.72,1.07] | 0.66 [0.47,0.92]* | 0.77 [0.57,1.05] |
| Three | 0.50 [0.34,0.73]*** | 0.71 [0.56,0.90]** | 0.50 [0.34,0.73]*** | 0.62 [0.43,0.89]** |
| Four or more | 0.48 [0.31,0.72]*** | 0.67 [0.52,0.87]** | 0.41 [0.27,0.62]*** | 0.57 [0.38,0.84]** |
| **Health insurance** | | | | |
| Not insured | 1 [Ref.] | 1 [Ref.] | 1 [Ref.] | 1 [Ref.] |
| Insured | 2.29 [1.65,3.18]*** | 1.40 [0.99,1.97] | 2.23 [1.59,3.11]*** | 2.26 [1.63,3.13]*** |
| **Wealth index** | | | | |
| Poorest | 1 [Ref.] | 1 [Ref.] | 1 [Ref.] | 1 [Ref.] |
| Poorer | 1.46 [1.06,2.02]* | 1.28 [1.00,1.64] | 1.20 [0.88,1.63] | 1.06 [0.79,1.43] |
| Middle | 1.56 [1.03,2.39]* | 1.31 [0.96,1.77] | 1.50 [0.98,2.28] | 1.28 [0.85,1.93] |
| Richer | 2.77 [1.60,4.79]*** | 1.48 [1.06,2.07]* | 2.85 [1.63,4.98]*** | 2.42 [1.43,4.08]** |
| Richest | 4.77 [2.13,10.66]*** | 2.25 [1.52,3.32]*** | 2.97 [1.37,6.45]** | 2.48 [1.27,4.86]** |
| **Place of residence** | | | | |
| Rural | 1 [Ref.] | 1 [Ref.] | 1 [Ref.] | 1 [Ref.] |
| Urban | 0.86 [0.64,1.16] | 1.13 [0.92,1.38] | 1.89 [1.39,2.57]*** | 1.45 [1.07,1.97]* |
| **Electricity** | | | | |
| No electricity | 1 [Ref.] | 1 [Ref.] | 1 [Ref.] | 1 [Ref.] |
| Has electricity | 1.40 [1.02,1.92]* | 1.20 [0.93,1.56] | 1.48 [1.09,2.03]* | 1.42 [1.03,1.95]* |
| **Region** | | | | |
| Western | 1 [Ref.] | 1 [Ref.] | 1 [Ref.] | 1 [Ref.] |
| Central | 0.63 [0.29,1.36] | 0.69 [0.42,1.14] | 0.82 [0.42,1.62] | 0.99 [0.48,2.05] |
| Greater Accra | 0.33 [0.15,0.72]** | 0.60 [0.37,0.99]* | 0.93 [0.43,2.05] | 1.21 [0.53,2.74] |
| Volta | 1.16 [0.47,2.82] | 0.58 [0.34,0.98]* | 2.62 [1.15,5.93]* | 1.37 [0.63,3.02] |
| Eastern | 0.61 [0.27,1.38] | 0.77 [0.47,1.29] | 1.71 [0.78,3.75] | 1.86 [0.81,4.26] |
| Ashanti | 0.70 [0.32,1.52] | 0.63 [0.39,1.01] | 2.07 [0.99,4.32] | 1.28 [0.61,2.68] |
| Western North | 0.59 [0.27,1.28] | 0.41 [0.24,0.69]*** | 2.15 [1.02,4.52]* | 1.21 [0.58,2.56] |
| Ahafo | 1.09 [0.49,2.42] | 0.88 [0.53,1.45] | 4.14 [1.86,9.22]*** | 4.32 [1.80,10.40]** |
| Bono | 0.77 [0.33,1.77] | 0.54 [0.32,0.91]* | 2.04 [0.93,4.50] | 2.49 [1.05,5.91]* |
| Bono East | 0.75 [0.36,1.59] | 0.67 [0.41,1.10] | 1.49 [0.75,2.94] | 1.22 [0.59,2.51] |
| Oti | 0.47 [0.23,0.98]* | 0.47 [0.28,0.78]** | 0.87 [0.45,1.67] | 0.99 [0.49,2.00] |
| Northern | 0.61 [0.29,1.29] | 0.39 [0.24,0.65]*** | 1.29 [0.66,2.52] | 0.76 [0.37,1.53] |
| Savannah | 0.64 [0.31,1.34] | 0.41 [0.25,0.68]*** | 1.23 [0.63,2.38] | 0.81 [0.40,1.65] |
| North East | 0.54 [0.26,1.14] | 0.43 [0.26,0.72]** | 1.77 [0.89,3.49] | 0.98 [0.48,2.01] |
| Upper East | 2.18 [0.90,5.31] | 0.81 [0.49,1.33] | 12.43 [4.66,33.19]*** | 4.59 [1.95,10.80]*** |
| Upper West | 1.83 [0.77,4.35] | 0.52 [0.31,0.86]* | 2.97 [1.42,6.22]** | 1.86 [0.87,3.99] |
| **Any barrier to healthcare** | | | | |
| No barrier | 1 [Ref.] | 1 [Ref.] | 1 [Ref.] | 1 [Ref.] |
| Any barrier | 0.82 [0.65,1.02] | 0.72 [0.63,0.84]*** | 0.87 [0.70,1.09] | 0.93 [0.75,1.15] |
| **Random Effects** | | | | |
| PSU variance | 0.4886 | 0.4389 | 0.5600 | 0.8301 |
| ICC | 12.9% | 11.8% | 14.5% | 20.2% |
| MOR | 1.948 | 1.881 | 2.042 | 2.385 |
| **Model Fitness** | | | | |
| Log-likelihood | -1565.23 | -2931.17 | -1603.66 | -1756.81 |
| AIC | 3224.47 | 5956.35 | 3301.32 | 3607.63 |
| Wald chi-square | 356.70*** | 332.83*** | 449.94*** | 294.46*** |
| N | 4,982 | 4,982 | 4,982 | 4,982 |
| Number of clusters | 617 | 617 | 617 | 617 |

aOR = adjusted odds ratio; 95% CI = 95% confidence intervals. * p<0.05  ** p<0.01  *** p<0.001. Ref. = Reference category. PSU = Primary Sampling Unit. ICC = Intra-Class Correlation. MOR = Median Odds Ratio. AIC = Akaike Information Criterion. Region entered as a 16-category factor (Western Region as reference) per convention in Ghana DHS multilevel analyses.
